# Divergent antibody-mediated population immunity to H5, H7 and H9 subtype potential pandemic influenza viruses

**DOI:** 10.1101/2025.09.08.25335309

**Authors:** Lucas Stolle, Jai S. Bolton, Rebecca Steventon, Reanna Gregory, Hannah Klim, Max Lee, Cecilia Di Genova, Luke Haddock, Aishwarya Bhatta, George Carnell, Caolann Brady, Jenny Redman, Jessica Hu, Kelly Da Costa, Matthew Edmans, Kate Calliss, Peter Simmonds, Edward M Hill, Nigel Temperton, Sarah C Hill, Lisa Jarvis, Carol McInally, Joe James, Ashley C. Banyard, Miles Carroll, Uri Obolski, Teresa Lambe, Alex Spencer, Anita Milicic, Nicole C Robb, Craig P Thompson

## Abstract

Influenza continues to cause significant mortality globally, and influenza viruses have substantial pandemic potential. Assessing pandemic risk requires a clear understanding of existing population immunity. Leveraging a unique large-scale cohort of human sera, we evaluated total and neutralising antibody-mediated immunity to multiple haemagglutinin (HA) proteins, including those from subtypes with high pandemic potential.

Our analysis reveals heterogeneous population immunity, with distinct age-dependent differences in neutralising responses to H5, H7, and H9 subtypes that align with historical circulation patterns of seasonal H1N1 and H3N2 viruses. Notably, H5 viruses were neutralised primarily via stem domain epitopes while H7 viruses were targeted mainly through head domain epitopes, although in both instances neutralisation via receptor binding site-adjacent epitopes was also detected. H5 responses were predominantly IgG1-mediated, whereas H7 responses were dominated by non-glycan-targeted IgG2 antibodies.

Cross-reactive antibodies to Group 2 potential pandemic HA had significantly lower avidity than those targeting closely related seasonal strains. These responses, generated through repeated seasonal influenza exposure, represent a broadly distributed but low-potency reservoir of immunity that could be boosted during a pandemic.

These findings highlight age-dependent variation in susceptibility to pandemic influenza and support vaccination approaches informed by exposure history.

## Introduction

The emergence and circulation of new influenza virus strains build population immunity. When individuals experience their first influenza infection in early childhood, specific B cell populations become established that can dominate future immune responses - a phenomenon known as ’antigenic imprinting’ [1]. Through repeated exposures to antigenically diverse influenza strains over time, the immune system progressively accumulates memory B cells capable of recognising conserved epitopes shared across multiple viral strains [2]. It is vital to assess the breadth and effectiveness of this accumulated population immunity, particularly considering the circulation of potential pandemic viruses for pandemic preparedness and vaccine development.

Currently, two influenza A subtypes, H1N1 and H3N2, co-circulate in the human population. H1N1 appeared in 1918 and circulated until 1957 when H2N2 replaced it. H2N2 then circulated until 1968 when H3N2 appeared. In 1977, H1N1 was reintroduced and circulated alongside H3N2 until 2009, when a novel H1N1 emerged (Supplementary Figure 1A). Presently, H3N2 dominates three out of every five influenza seasons [3].

Alongside these human-adapted viruses, there is the constant circulation of avian influenza strains amongst wild and domestic birds and other animals. Of particular concern in recent years is the circulation of highly pathogenic H5Nx 2.3.4.4b lineages (primarily H5N1) of influenza due to its unprecedented global spread, high pathogenicity and an increased ability to infect mammals [4]. Likewise, H7N9 has caused hundreds of human infections in China since 2013 with a case fatality rate of around 40%. H9N2 viruses circulate in poultry worldwide and have repeatedly infected humans, though typically causing milder illness than H5 or H7 viruses. Consequently, understanding the extent (if any) and form of population immunity to potential pandemic viruses is of high importance to inform public health initiatives [5].

Influenza A virus is primarily an avian virus that crosses into humans via zoonotic transfer, which can lead to pandemics. Influenza haemagglutinins (HAs) are divided into 19 antigenic subtypes (H1-H19), with H1, H2, H5, and H9 classified phylogenetically as ’Group 1’, while H3 and H7 belong to ’Group 2’ (Supplementary Figure 1B). Neutralisation occurs through antibodies binding either to the receptor binding site (RBS) within the HA head domain or to the HA stem domain preventing membrane fusion. In blood, IgG is the dominant antibody class, consisting of subclasses IgG1-4, with IgG1 providing the greatest neutralisation capacity, while IgG3’s larger hinge domain may enable neutralisation of a wider array of drifted influenza strains [6].

Reported studies of population immunity to the 2.3.4.4b lineage of H5N1 influenza show considerable variation with some studies using live virus neutralisation assays to detect the presence of neutralising antibodies in 20% of individuals screened [4], whilst other groups using the same assay, unable to detect any neutralisation [7, 8]. Other studies have demonstrated that seasonal influenza vaccines can elicit cross-reactive antibody responses against avian H5N1, and to a lesser extent, H7N9 viruses [9–12]. The basis for this variation in detected immunity remains unclear.

The mechanistic basis for this cross-protective immunity has been proposed to be primarily mediated by antibodies directed against highly conserved epitopes within the HA stem domain, which exhibits structural conservation across related influenza subtypes [13].

Here, we assess and characterise population immunity to seasonal and potential pandemic influenza A viruses using a pseudotyped influenza virus microneutralisation assay (pMN) in a Scottish blood donor cohort collected in 2020. Collectively, we have run 20,182 neutralisation assays, producing 4,646 data points, using cohorts of 195-512 blood donors against 19 pseudotyped influenza viruses spanning over 100 years of influenza virus evolution, to produce a comprehensive representation of antibody-mediated immunity to influenza A in the human population (Supplementary Table 1).

The study reveals age-dependent shifts in neutralising immunity to H5/H9 (Group 1) and H7 (Group 2) subtype viruses that correspond to historical H1N1 and H3N2 circulation patterns, explaining previously observed demographic and epidemiological trends of H5/H7-associated mortality and incidence [14]. Flow cytometry showed distinct IgG subclass binding patterns, with IgG2 preferentially binding H7 HA and IgG1 binding H5 HA. Notably, we find that H5 neutralising immunity is stem domain dominant while H7 immunity is head domain dominant. However, in both instances, we detected cross-reactive neutralisation that maps to RBS epitopes. We demonstrate that cross-reactive antibodies to potential pandemic viruses are of lower avidity than those targeting seasonal strains, providing a mechanistic explanation for the variable detection of population immunity across assay platforms.

## Results

### Identification of divergent age-dependent neutralising antibody population immunity profiles to H5/H9 and H7 subtype potential pandemic influenza viruses

We assessed population immunity using pseudotyped viruses displaying HAs from a chronologically distinct set of influenza viruses, tested against blood donor sera collected in the UK in 2020 from individuals aged 18-70 years.

Our pseudotyped virus panel consisted of 11 seasonal H1 and H3 viruses, a previously circulating H2 strain, and 6 potential pandemic H5, H7, and H9 viruses, including one virus from the 2.3.4.4b lineage (Supplementary Table 1) to enable us to assess pan-subtype cross-reactivity resulting from H1N1 and H3N2 circulation.

To ensure the validity of our data, we established the neutralisation threshold as five standard deviations above the mean background neutralisation of an Ebola pseudotyped virus, which Scottish blood donors are not expected to neutralise (Supplementary Figure 2A, n=100) [15]. We also performed receptor-destroying enzyme (RDE) assays and used mismatched ferret antisera as controls (Supplementary Figure 2).

We detected substantial neutralising antibody-mediated population immunity to H5 and H9 viruses via pseudotype microneutralisation assay (pMN), including the A/mute_swan/England/117298/2022 2.3.4.4b lineage H5 virus, close relatives of which have recently infected several humans and are circulating widely in poultry and cattle. All these pseudotyped viruses showed broad population reactivity, with 90% or more of sera samples demonstrating neutralising activity. In contrast, population immunity to H7 pseudotyped viruses was lower, with detectable immune responses present in only 55-65% of samples. (Supplementary Figure 3).

This trend continued in terms of the magnitude of neutralisation response with three of five seasonal H1 viruses demonstrating higher median titres than H5 viruses, while all seasonal H3 viruses exceeded H7 virus median titres (Supplementary Figure 3).

A notable exception to the broad Group 1 reactivity was A/England/1/1966 H2N2, which was neutralised by only 23.7% of samples.

Age-dependent neutralisation responses were visualised using locally weighted scatterplot smoothing (LOWESS) curves with a smoothing parameter of 0.4. Bootstrapped 95% confidence intervals were calculated for each curve (Supplementary Figure 5).

Age-dependent responses revealed striking patterns that differed markedly between viruses. Group 1 H5 and H9 neutralisation responses increased with age, reaching peak levels in individuals born before 1957 (Figure 1A, Supplementary Figures 5 & 6)). For example, peak neutralisation for H5 A/mute_swan/England/117298/2022 2.3.4.4b and H5 A/Vietnam/1203/2004 viruses as well as H9 A/Hong Kong/308/2014 pseudotyped viruses, occurred in individuals born before 1957.

**Figure 1:**
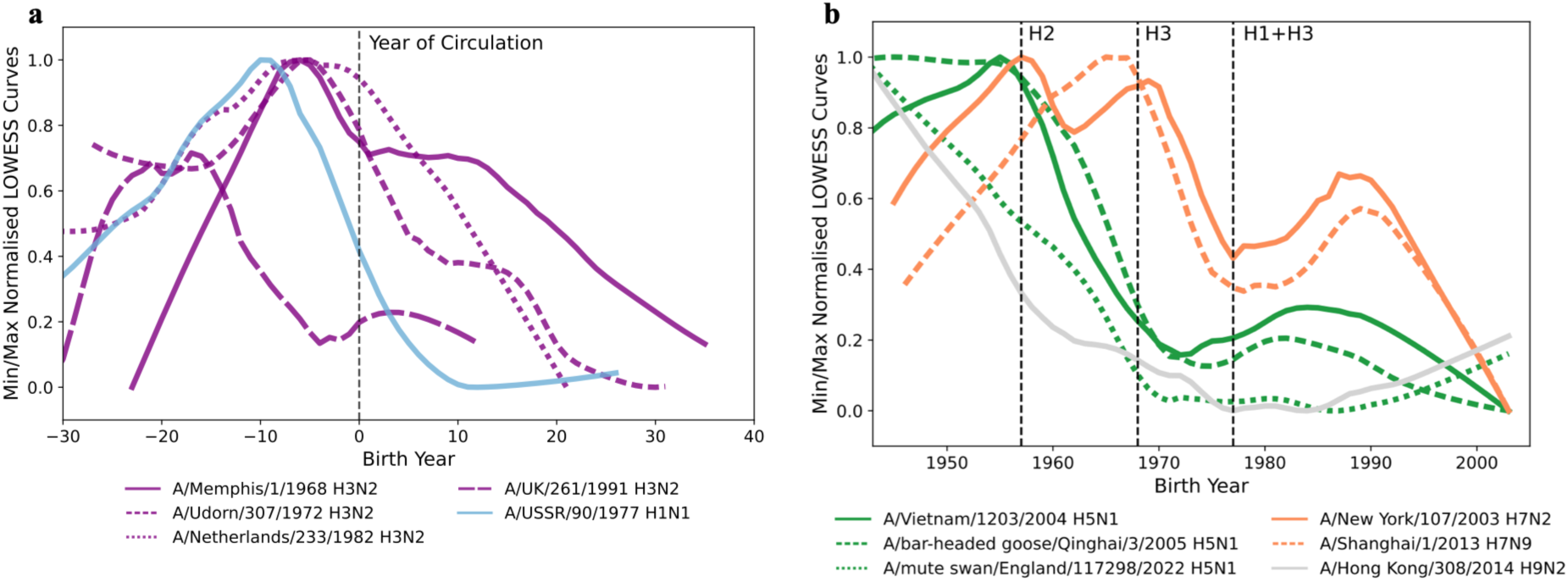
Antibody-mediated immune profiles to seasonal and potential pandemic influenza strains are age-and subtype-dependent. **a.** LOWESS curves denoting the imprinting of seasonal influenza viruses, where the difference between birth year and year of virus circulation is calculated on the x axis, and the vertical dashed line indicates individuals born in the year the virus emerged. Peaks typically align in those born 6 years prior to virus circulation, although A/UK/261/1991 diverges from this trend. **b.** LOWESS curves denoting imprinting patterns for Group 1 vs Group 2 potential pandemic pseudotyped viruses, showing divergent age-dependent patterns. The vertical dashed lines indicate when the respective subtypes denoted at the top of the lines began circulating in humans, replacing the previous subtype. Sample sizes can be found in Table S1. LOWESS curves were fitted with a smoothing parameter of 0.4. Bootstrapped 95% confidence intervals are shown in Supplementary Figure 4. Source data are provided as a Source Data file.

Unexpectedly, H7 peak neutralisation responses were shifted towards individuals born shortly before H3N2 emergence in 1968. For example, peak neutralisation of H7 A/Shanghai/1/2013 occurred in individuals born around 1965, with similar timing observed for H7 A/New York/107/2003 (Figure 1A).

These data suggest that there may be low levels of cross-reactive antibodies and associated B cells in the human population to potential pandemic H5, H7 and H9 influenza viruses, which could be boosted during an outbreak.

Seasonal virus neutralisation patterns were also indicative of imprinting (Figure S4). For seasonal H1N1 and H3N2 viruses that emerged during birth years relevant to our sample cohort, neutralising responses peaked in individuals born approximately 6 years before virus emergence (Figure 1B), supporting the concept of immunological imprinting during early childhood.

Intriguingly, we found that the age-dependent response for a subset of viruses broke this trend, with maximum responses that do not follow this typical imprinting pattern. For example, the H3 A/UK/261/1991 virus has atypical peaks at 15-25 and 45 years pre-circulation (Figure 1B).

To elucidate any relationships between an individual’s neutralising responses to two different viruses, we calculated Spearman’s correlation coefficients (Supplementary Figure 7). Almost all of the strongest correlations (ρ=0.4 - 0.81), were clustered among the H5 and H9 potential pandemic subtypes, likely due to shared or structurally similar antigenic epitopes.

Surprisingly, the recently circulated A/Brisbane/2/2018 H1N1 and the more distantly circulated A/England/1/1966 H2N2 viruses also exhibited a strong correlation (ρ=0.75).

### Differential IgG subclass responses occur towards different potential pandemic influenza HAs

To determine which IgG subclasses were involved in the observed cross-reactivity between historical, seasonal and potential pandemic influenza HAs, flow cytometry was performed using sera samples with high levels of neutralising antibodies.

IgG1-4 were detectable against all tested viruses (Figure 2) with antibody specificity validated via monoclonal antibody controls (Supplementary figure S8). In samples that were identified as having high levels of H5 pseudotyped virus neutralisation, IgG1 binding was higher to potential pandemic virus H5 HAs from A/Vietnam/1203/2004 and 2.3.4.4b lineage H5N1 virus A/Texas/37/2024 than H1 seasonal virus HAs A/Brisbane/2/2018 and A/USSR/90/1977 (VN vs USSR, p= ≤0.0001, Texas vs USSR p= 0.0138, Brisbane vs VN p = 0.0011). IgG2 levels bound the seasonal H1 HAs to a greater extent than the potential pandemic H5 HAs (USSR vs VN, p= 0.0022, USSR vs Texas, p= 0.0001, USSR vs Brisbane, p= 0.0004). IgG3 and IgG4 were at low levels, and so no conclusions could be drawn (Supplementary Figure 9).

**Figure 2.**
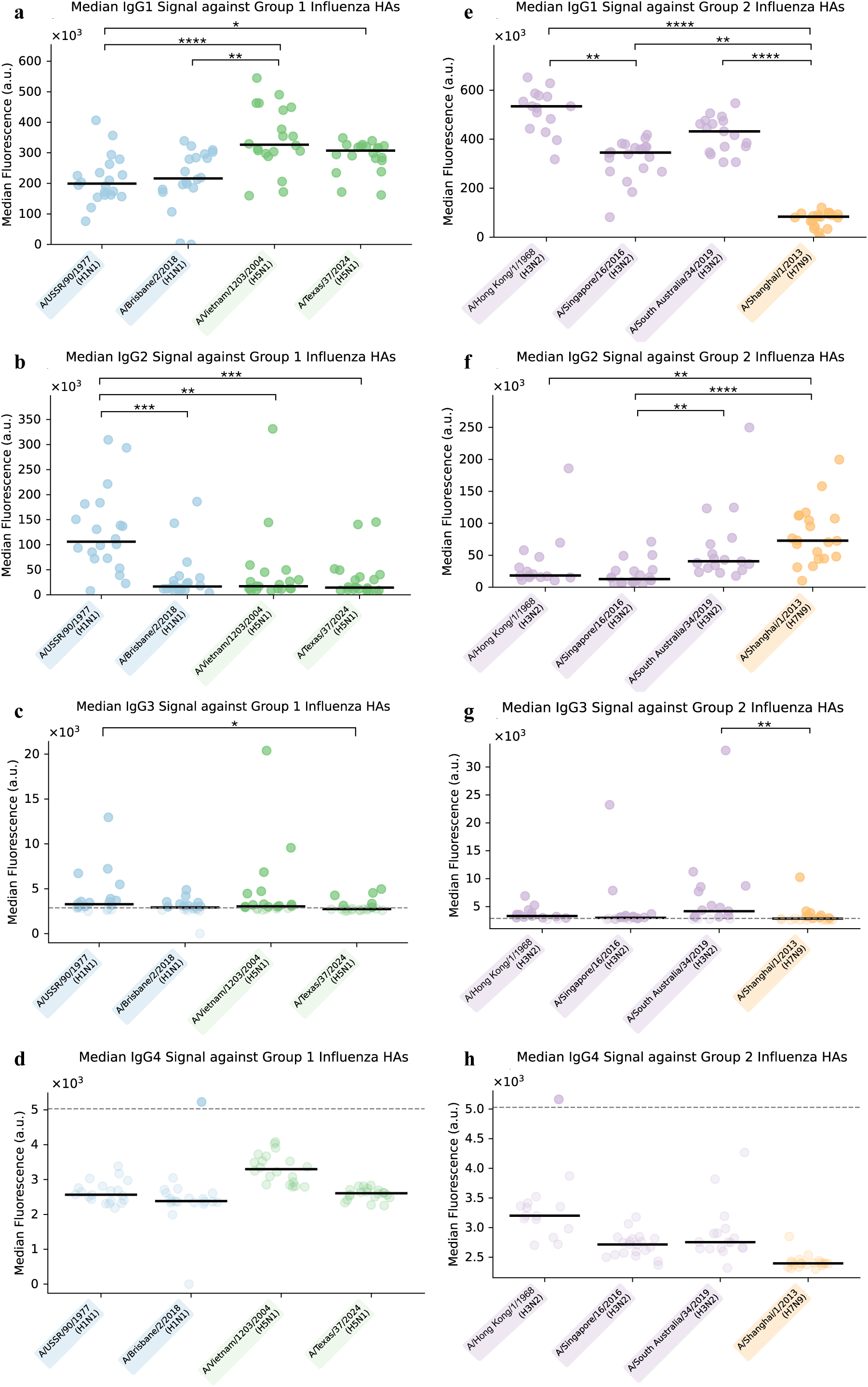
Different IgG subclasses preferentially bind to potential pandemic influenza HAs. Flow cytometry analysis of IgG subclasses bound to Group 1 and Group 2 full length influenza haemagglutinin (HA). **a-d.** IgG1-4 composition of Group 1 neutralizing samples run against seasonal H1 – A/USSR/90/1977 and A/Brisbane/2/2018 – and H5 HAs – A/Vietnam/1203/2004 and 2.3.4.4b lineage A/Texas/37/2024. **e-h.** IgG1-4 composition of Group 2 neutralizing samples specifically H3N2 HAs – A/Hong Kong/1/1968, A/Singapore/16/2016, A/South Australia/34/2019 – and H7N9 HAs – A/Shanghai/1/2013. This data shows that antibodies in the human population bind to potential pandemic influenza HAs. Notably the greatest proportion of Group 1 virus IgG1 binds to potential pandemic H5N1 HAs, A/Vietnam/1203/2004 and 2.3.4.4b lineage A/Texas/34/2024, rather than seasonal H1 HAs, A/USSR/90/1977 or A/Brisbane/2/2018. N = 30 individual donor sera. Horizontal line indicate medians. Statistical analysis was performed using two-tailed Kruskal-Wallis tests with Dunn’s correction at a 95% confidence level. Asterisks denote statistical significance as recommended by GraphPad Prism: p < 0.032 (*), p < 0.0021 (**), p < 0.0002 (***), p < 0.0001 (****). Only statistically significant p-values are depicted. Dashed line represents the limit of detection based on an IgG1-4 standard. Median fluorescence is expressed in arbitrary units (a.u.). Exact p-values are provided in the Source Data file. Source data are provided as a Source Data file.

In contrast to the H1 and H5 subtype (Group 1) viruses, H3 and H7 (Group 2) viruses demonstrated the opposite relationship regarding IgG1 and IgG2. Seasonal H3 virus HAs from A/Hong Kong/1/1968, A/Singapore/16/2016 and A/South Australia/34/2019 bound significantly more IgG1 than the potential pandemic H7 HA from A/Shanghai/1/2013 (Figure 2; Shanghai vs SA p= ≤0.0001, Shanghai vs HK p=≤0.0001, Shanghai vs Singapore p = 0.0035). However, significantly more IgG2 bound the A/Shanghai/1/2013 HA than seasonal H3 HAs (Figure 2; Shang vs HK p=0.006, Shanghai vs Singapore p= ≤0.0001).

As IgG2 is associated with binding to polysaccharide antigens [16], we sought to assess whether the greater IgG2 binding to the A/Shanghai/1/2013 HA was due to glycosylation. We therefore deglycosylated the H7 A/Shanghai/1/2013 HA as well as the H3 HA from A/Singapore/16/2016 via PNGase F treatment. Deglycosylation of the HAs significantly increased IgG2 binding in both instances when assessed via flow cytometry (Supplementary Figure 10, D, p=<0.0001). To determine the impact of deglycosylation on neutralisation, we deglycosylated the H7 A/Shanghai/1/2013 pseudotyped influenza virus. The deglycosylated H7 A/Shanghai/1/2013 pseudotyped influenza virus did not significantly differ in neutralisation to the non-deglycosylated A/Shanghai/1/2013 pseudotyped influenza virus (Supplementary Figure 10B). In both instances, deglycosylation was confirmed by western blot or SDS-PAGE (Supplementary Figure 10A & E).

To investigate whether the distinct IgG subclass responses reflected differences in epitope targeting, we compared IgG1 and IgG2 binding to full-length HA ectodomain and HA1 (head domain only) constructs of A/Shanghai/1/2013 (H7N9). IgG1 binding was markedly reduced when using the HA1 construct compared with the full-length HA (8.0-fold reduction in median GMFI, Wilcoxon *p*<0.0001, adjusted *p*<0.001), indicating that the majority of the IgG1 response recognises stem-domain epitopes (Supplementary Figure S14a). In contrast, IgG2 binding did not differ significantly between the HA1 and full-length constructs (1.1-fold difference, Wilcoxon *p*=0.093, adjusted *p*=0.19), supporting preferential recognition of head-domain epitopes by IgG2 antibodies (Supplementary Figure S14b). These findings provide a mechanistic explanation for the differential IgG subclass binding observed across influenza HA subtypes.

In summary, our flow cytometry analysis revealed distinct IgG subclass profiles towards potential pandemic influenza HAs. IgG1 responses preferentially recognised H5 HAs over seasonal H1 HAs, whereas IgG2 responses were enriched against H7 HA compared to seasonal H3 HAs. Glycosylation partially masked IgG2-binding epitopes but did not alter H7 neutralisation, while comparison of full-length and HA1 constructs demonstrated that IgG1 predominantly targets stem-domain epitopes and IgG2 preferentially recognises head-domain epitopes. Together, these findings indicate that influenza HA domain specificity differs between IgG subclasses and may contribute to subtype-specific patterns of antibody-mediated immunity.

### H5 virus neutralisation is stem domain mediated whilst H7 virus neutralisation is head domain mediated

Previous studies have identified via ELISA that immunity to the 2.3.4.4b lineage A/Texas/37/2024 H5 HA is largely stem-mediated [17]. To assess whether this stem mediated immunity translated into neutralisation via pMN assay and to further characterise the neutralising immune response to potential pandemic viruses, we performed antibody pulldowns using both the full-length (ectodomains) and head domain proteins.

Purified antibodies against the full-length proteins and head domains of A/Texas/37/2024 were assessed via pMN assay against A/USSR/90/1977, H5 A/Vietnam/1203/2004 and H5 A/Texas/37/2024 pseudotyped viruses. In each instance, sera samples identified as having high levels of neutralising antibodies to the antigen were used in the pulldown.

For the 2.3.4.4b lineage H5 A/Texas/37/2024 antibody pulldowns, we found that neutralisation of historical H1 strains and potential pandemic H5 pseudotyped influenza viruses was greater when the full-length protein was used rather than the head domain protein, indicating substantial stem domain cross-reactive antibodies (Figure 3A, p=0.00043) in line with previously published studies [20, 41] However, head domain antibodies cross-reactive against A/Vietnam/1203/2004 and A/Texas/37/2024 H5 viruses were also detected in a small number of samples.

**Figure 3.**
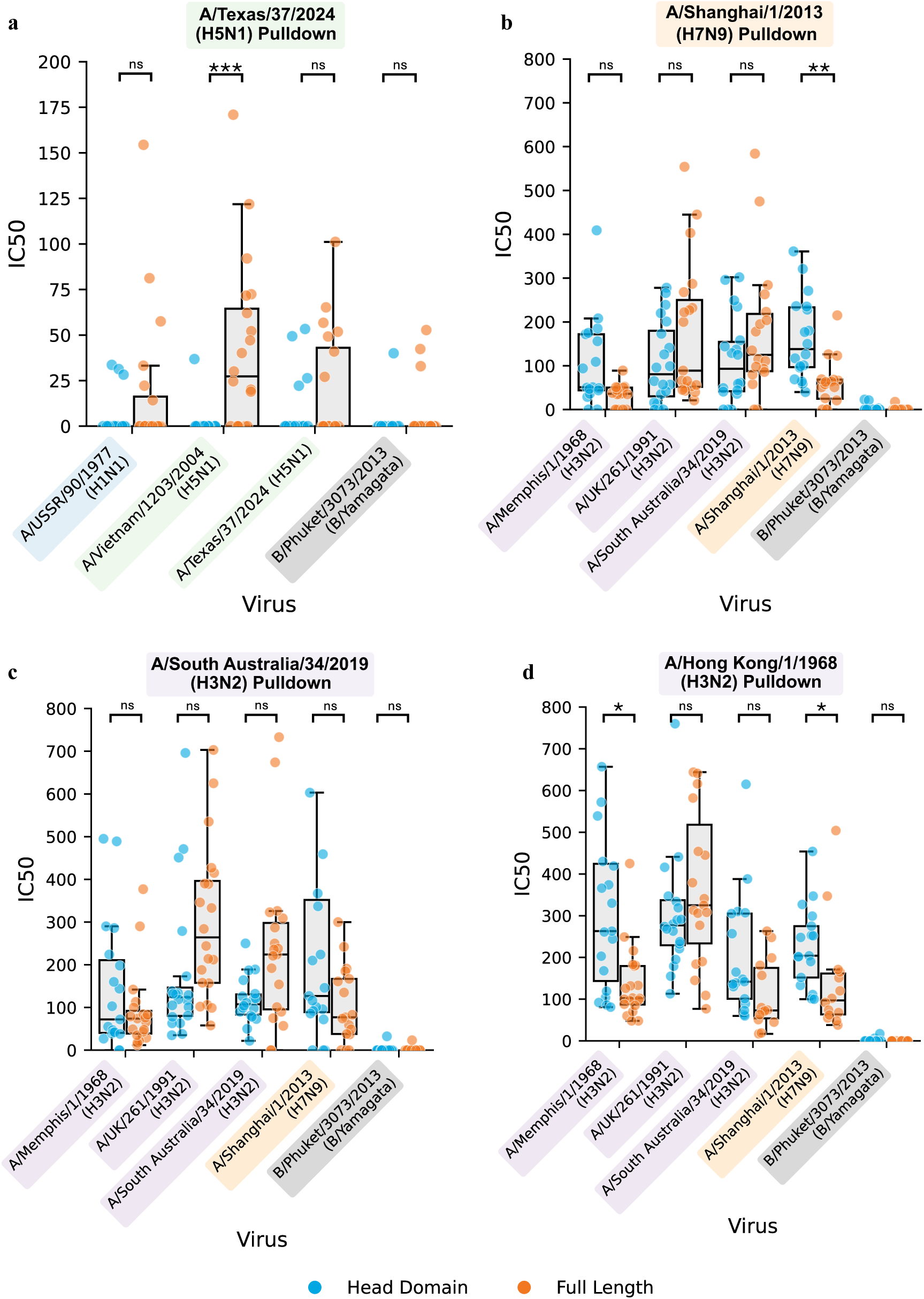
Head versus stem domain-mediated neutralisation differs between Group 1 and Group 2 influenza haemagglutinins. Antibody pulldown assays were performed using both head domain (blue) and full-length HA proteins (orange) against a subset of pseudotyped influenza viruses. **a.** Pulldown assays using full-length and head domain constructs of H5 HA from A/Texas/37/2024 were tested against a panel of H1 and H5 pseudotyped viruses. **b.** Pulldown assays using full-length and head domain constructs of H7 HA from A/Shanghai/1/2013 were tested against a panel of H3 and H7 pseudotyped viruses. **c.** Pulldown assays using full-length and head domain constructs of H3 HA from A/South Australia/34/2019 were tested against a panel of H3 and H7 pseudotyped viruses. **d.** Pulldown assays using full-length and head domain constructs of H3 HA from A/Hong Kong/1/1968 were tested against a panel of H3 and H7 pseudotyped viruses. In all experiments, B/Phuket/3073/2013 served as a negative control. N = 20 individual donor sera. Y-axis denotes neutralisation in 50% inhibitory concentration (IC50) units. Box plots show the median (centre line), the 25th and 75th percentiles (box bounds), and whiskers extending to the most extreme data points within 1.5× the interquartile range; all individual data points are overlaid, showing the minima and maxima. Statistical analysis was performed using two-tailed Mann-Whitney U tests with Holm-Bonferroni correction at a 95% confidence level. Asterisks denote statistical significance: p < 0.05 (*), p < 0.01 (**), p < 0.001 (***). Exact p-values are provided in the Source Data file. Source data are provided as a Source Data file.

To assess whether this trend extended to H3 and H7 viruses, we performed antibody pulldowns using the full-length and head domain proteins of H3N2 viruses A/Hong Kong/1/1968, A/South Australia/34/2019, as well as H7 virus A/Shanghai/1/2013. The antibody pulldowns were run against H3 pseudotyped viruses A/Memphis/1/1968, A/UK/261/1991, A/South Australia/34/2019 and H7 pseudotyped virus A/Shanghai/1/2013 (Figure 3B-D).

In contrast to H1 and H5 antibody-mediated population immunity, H3 and H7 antibody-mediated immunity was mainly mediated by head domain targeting antibodies (Figure 3B-D). For the seasonal H3 A/Hong Kong/1/1968 and potential pandemic H7 A/Shanghai/1/2013 pulldowns, head-mediated neutralisation was significantly greater than neutralisation conferred by the respective full-length proteins (Shanghai, p=0.00358, Hong Kong p=0.01). No significant differences were identified for the seasonal H3 A/South Australia/34/2019 pulldown (Figure 3B & D).

Consequently, this data demonstrates that antibody-mediated population immunity to potential pandemic H5 viruses is mainly stem-mediated, with the presence of head domain antibodies at a significantly lower level. In contrast, immunity to potential pandemic H7 viruses is mainly head domain mediated.

### Head domain epitopes located around the receptor binding domain mediate population immunity to potential pandemic influenza

Our assessment of head and stem-mediated immunity for potential pandemic H5 and H7 viruses demonstrated the presence of head domain mediated antibodies able to bind both H5 and H7 HAs and neutralise the respective pseudotyped influenza viruses (Figure 3). To further characterise the head-mediated cross-reactive antibody responses identified between H1 and H5, as well as H3 and H7, we used structural bioinformatics to map potentially cross-reactive antibody binding sites of low variability. We have used this approach previously to successfully identify cross-reactive H1N1 epitopes [18].

The approach identified several regions of potential conservation in the different HAs. To determine if these predicted antibody binding sites were responsible for the observed cross-reactivity, site-directed mutagenesis (SDM) was carried out on key residues. All key residues selected for mutation were chosen via alignment of current and historical HAs sequences to identify charged residues which moved through a limited repertoire of identities as influenza strains evolved over time.

In all instances, broadly neutralising stem-targeted antibodies were used as controls to normalise the neutralisation of wild-type and mutant pseudotyped viruses. Furthermore, several antibody binding sites that appeared to be variable in the bioinformatic analysis were used as controls.

For H1/H5, bioinformatic analysis revealed two low variability putative antibody binding sites in the head domain of H5 – positions S145 and T215 (H5 numbering) (Figure 4A). Position 145 is a structural analogue of position 147 in H1 (133 H3 numbering), part of an epitope of low variability identified by Thompson et al. [18] that is close to the RBS. S145 was mutated to leucine in the H5 A/bar-headed goose/Qinghai/3/2005 background, which significantly reduced neutralisation in human sera compared to the control mAb (145L p=0.0012, Figure 4D). A similar response was also seen when position 215 was mutated (215K p=0.0159, 215L p=0.0028). Other control mutations were made in the H5 head domain; D281 and D70 (Figure 4D), which elicited no statistically significant effects on neutralisation.

**Figure 4.**
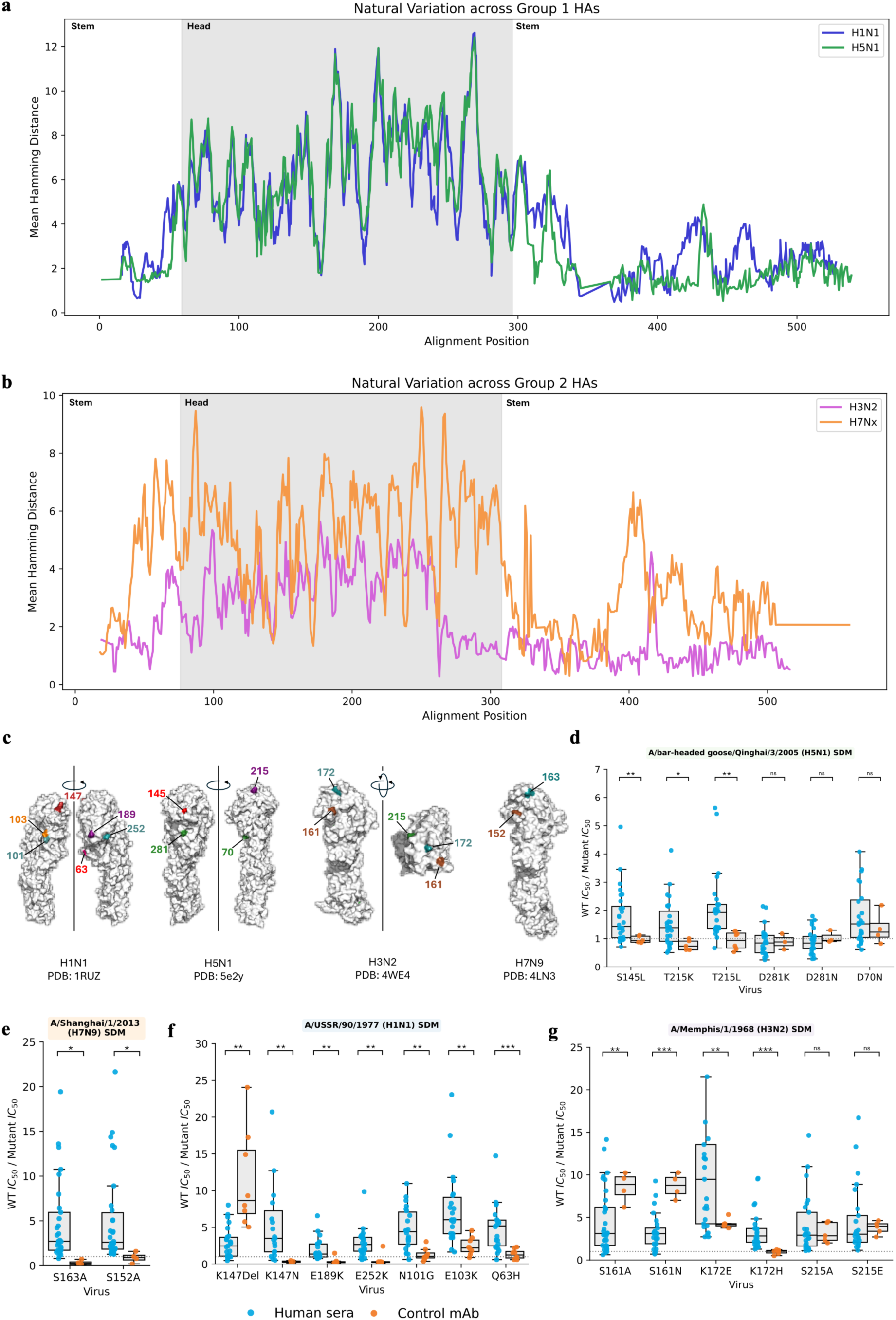
Epitopes around the receptor binding site mediate neutralisation to potential pandemic strains. **a-b.** Variability of predicted antibody binding sites on the surface of the H1 C H5 HAs (**a.**) as well as the H3 C 7 HAs (**b.**). x-axis denotes the variability of the central amino acid position with an 800A^2^ antibody binding site. Shared area denotes the head domain of HA, whilst non-shaded area denotes the stem domain. **c.** Crystal structures of the H1, H5, H3 and H7 HA monomers with associated pdb numbers showing the position of amino acids mutated in **d-g**. **d.** Amino acid mutations in the A/bar-head goose/Qinghai/3/2005 HA. **e.** Mutations in the A/Shanghai/1/2013 background. **f.** Mutations in the A/USSR/90/1977 background. **g.** Mutations in A/Memphis/1/1968 background. CR6261 was used as a control monoclonal antibody for Group 1, and CR8020 was used for Group 2. Y-axis denotes neutralisation in 50% inhibitory concentration (IC50) units. N = 25 individual donor sera. Box plots show the median (centre line), the 25th and 75th percentiles (box bounds), and whiskers extending to the most extreme data points within 1.5× the interquartile range; all individual data points are overlaid, showing the minima and maxima. Statistical analysis was performed using two-tailed Student’s t-tests with Holm-Bonferroni correction on the log_10_ of the ratios at a 95% confidence level. Asterisks denote statistical significance: p < 0.05 (*), p < 0.01 (**), p < 0.001 (***). Exact p-values are provided in the Source Data file. Source data are provided as a Source Data file.

Consequently, the SDM revealed the existence of head-targeting antibodies against H5 and shared epitopes in the head domains of H1 and H5.

Similar trends in neutralisation reduction were also identified for the corresponding mutations in the H1 A/USSR/90/1977 HA head domain (Figure 4F). Mutation of 147 in H1 (S145 in H5 and 133 in H3 numbering) significantly decreased neutralisation of A/USSR/90/1977 (147Del p<0.001, 147N p=0.0029). However, in this instance, all mutations made in the head domain caused statistically significant reductions in neutralisation.

To validate our approach, we used sera from a population of mute swans (*Cygnus olor*) that was exposed to multiple waves of naturally occurring H5 HPAIV. These swan samples were collected between June 2017 and July 2019. As expected, most birds tested showed some evidence of infection with H5 subtype influenza during this period, making them ideal to contrast with the cross-reactive antibody profiles produced in human samples from seasonal influenza infection [19, 20].

Remarkably, these samples produced a similar pattern of neutralisation reduction at the majority of the same residues; reduced neutralisation for S145L, S145A, T215L (p<0.001, p=0.0035, p=0.04061) and no reduction for D281K and D70N. Again, mutations that reduced neutralisation were found around the RBS of the H5 HA, which mirrored those identified with human sera (Supplementary Figure 12B).

The same approach was applied to H3 and H7 HAs. Key residues in putative antibody binding sites containing regions of lower variability were mutated in the H3 A/Memphis/1/1968 and H7 A/Shanghai/1/2013 backgrounds. Control mutations and CR8020, a monoclonal antibody targeting the stem, were used for normalisation in each instance.

Positions S152 and S163 (H7 numbering) were identified as residues that may fall within shared, relatively conserved antibody binding sites for H3 and H7 (Figure 4B). Mutation of positions S152 or S163 in the H7 A/Shanghai/1/2013 background showed a significant reduction in neutralisation (p=0.0298, p=0.03241, Figure 4E). Corresponding mutations were made in the H3 A/Memphis/1/1968 background, where a significant reduction in neutralisation was also seen for position S163 (K172 in H3, 172E p=0.0025, 172H p<0.001). However, an increase in neutralisation was seen when position S152 (S161 in H3, 161A p=0.0013, 161N p<0.001) was mutated. A control mutation was made at position 215, which showed no change in neutralisation (Figure 4G).

Collectively, these data demonstrate that human sera can neutralise non-circulating potential pandemic H5 and H7 HA pseudotyped influenza viruses via head domain epitopes

### Cross-reactive antibodies to potential pandemic viruses are of lower avidity than those targeting seasonal strains

To investigate why studies report variable levels of population immunity to potential pandemic viruses using live virus assays [4], we compared pseudotyped and live virus neutralisation for a panel of H3N2 viruses and assessed antibody avidity across subtypes.

We performed live virus microneutralisation assays using H3N2 viruses A/X-31 (H3N2 1968 HA and NA with PR8 core), A/Udorn/307/1972, A/X-79 (H3N2 1982 HA and NA with PR8 core), and A/Caen/1/1984 (H3N2) against a subset of blood donor sera (n=29) that had been previously assessed by pMN. Pseudotyped virus neutralisation was detectable in most samples for each virus. However, live virus neutralisation was restricted to a smaller subset, typically peaking in individuals who had been born +/-5 years of virus circulation, with many pseudotype-positive samples showing no detectable live virus neutralisation (Figure 5 and Supplementary Figure 13).

**Figure 5.**
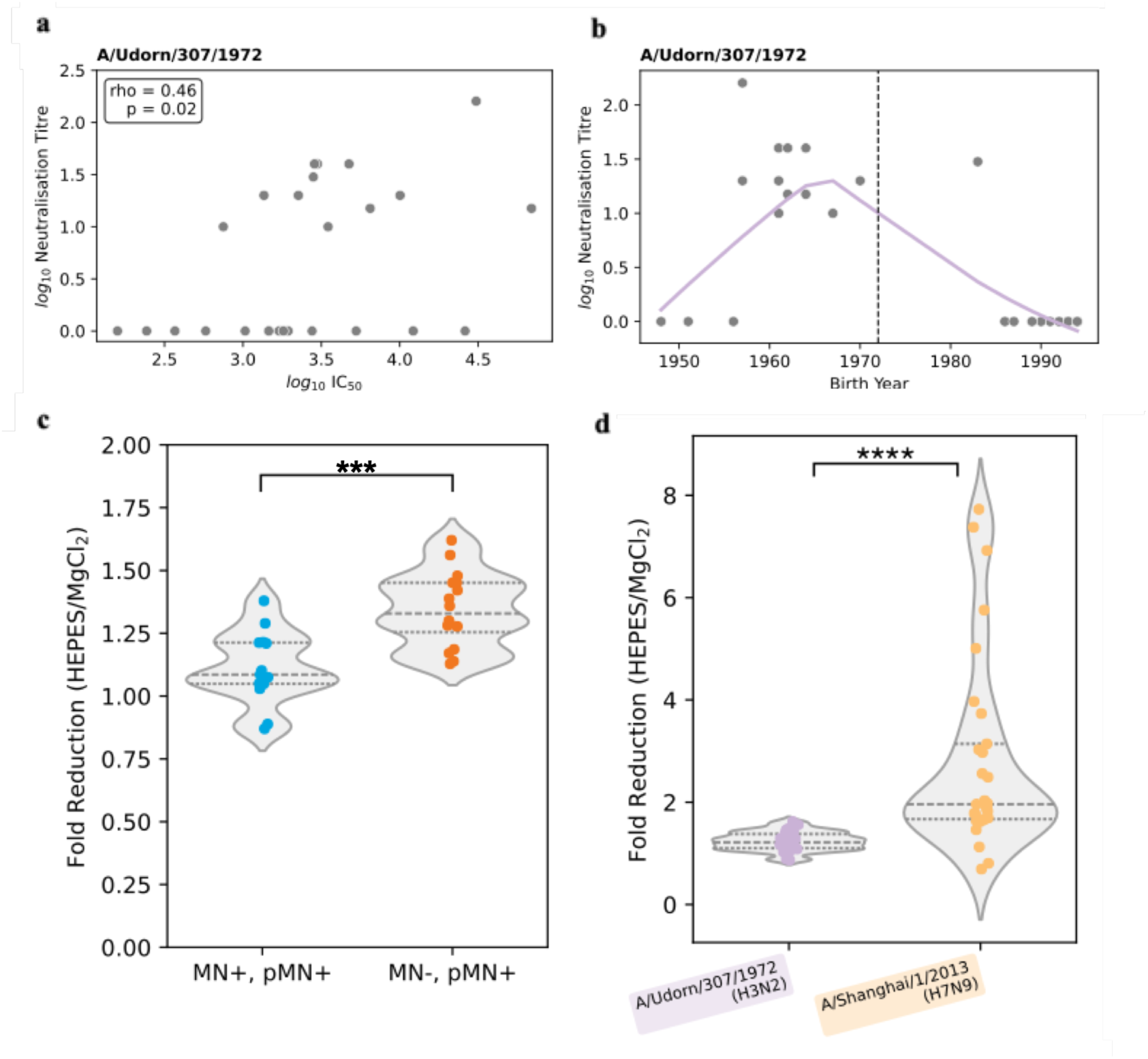
Avidity of cross-reactive antibodies and live virus microneutralisation concordance. a. Pseudotype microneutralisation (pMN) IC50 plotted against live virus microneutralisation titre (MN, neutralisation titre) for A/Udorn/307/1972 (H3N2). Two-tailed Spearman’s correlation coefficient and p-value indicated. b. Live virus microneutralisation titres plotted against birth year for A/Udorn/307/1972. The vertical dashed line indicates the year the virus began circulating in humans. LOWESS trendline fitted with a smoothing parameter of 0.4. c. Avidity index comparison between samples positive by both pMN and live virus MN (pMN+/MN+) and samples positive by pMN only (pMN+/MN−) for A/Udorn/307/1972. Statistical analysis was performed using a two-tailed Mann-Whitney U test. d. Avidity index comparison between A/Udorn/307/1972 (H3N2) pMN-positive and A/Shanghai/1/2013 (H7N9) pMN-positive samples, showing significantly lower avidity for H7 cross-reactive antibodies. Statistical analysis was performed using a two-tailed Mann-Whitney U test. N = 29 for all panels. Asterisks denote statistical significance: p < 0.05 (*), p < 0.01 (**), p < 0.001 (***), p < 0.0001 (****). Exact p-values are provided in the Source Data file. Source data are provided as a Source Data file.

Pseudotyped and live virus neutralisation titres were weakly or not correlated for three of the four viruses tested (X-31: Spearman ρ=0.20, p=0.3; X-79: ρ=0.23, p=0.24; Caen/84: ρ=0.24, p=0.22), while A/Udorn/307/1972 showed a moderate but statistically significant correlation (ρ=0.40, p=0.033), indicating that the assays partially agree but with substantial discordance driven by samples that are pseudotype-positive but live-virus-negative (Supplementary Figure 14). Assay validity was confirmed by internal controls and positive control sera.

This discordance was more pronounced for potential pandemic viruses. Live virus neutralisation assays were performed against A/chicken/England/014330/2022 (H5N1 2.3.4.4b) and A/chicken/England/1158-11406-1/2008 (H7N7) using the same sera. No detectable neutralisation was observed from blood donor samples for either virus, despite broad pseudotype neutralisation of H5 and H7 viruses in these same individuals. Assay validity was again confirmed using infected animal sera and internal controls (Supplementary Table 2 and Supplementary Figure 14).

To determine whether antibody avidity could explain the discordance between assays, we performed avidity ELISAs using MgCl₂ chaotropic washing against the H3 A/Udorn/307/1972 HA. Sera were stratified by their live virus neutralisation result: samples positive in both pseudotype and live virus assays versus those positive in the pseudotype assay alone. Samples that were pseudotype-positive but live-virus-negative had significantly lower antibody avidity than those positive in both assays (p<0.001, Figure 5), demonstrating that antibody avidity determines whether cross-reactive neutralisation is detectable by live virus assay.

To assess whether avidity differed across subtypes, we compared antibody avidity against the H3 A/Udorn/307/1972 and H7 A/Shanghai/1/2013 HAs. Antibodies binding the H7 HA showed significantly lower avidity than those binding the H3 HA (p<0.0001, Figure 5), consistent with H7 cross-reactive antibodies being generated at greater antigenic distance from the immunising seasonal virus. In support of this model, pseudotype and live virus neutralisation titres showed strong concordance for contemporary influenza B viruses where high-avidity antibodies predominate [43]. This avidity gradient provides a mechanistic explanation for the lower detection of H7 and H5 population immunity by live virus assays compared to pseudotype-based approaches.

## Discussion

We identified age-dependent antibody responses to pandemic H5, H7, and H9 viruses with distinct IgG subclass patterns: IgG1 preferentially bound H5 over seasonal H1 HAs, while IgG2 bound H7 over H3 HAs. H1/H5 subtype viruses (Group 1) elicited primarily stem-targeting antibodies, whereas H3/H7 subtypes (Group 2) primarily induced head domain targeting antibodies. However, in both instances, head domain antibodies were identified and mapped for both H1/H5 and H3/H7 subtypes that targeted RBS epitopes.

We found that 93-100% of samples assessed exhibited neutralisation against H5 subtype A/Vietnam/1203/2004, A/bar-headed goose/Qinghai/3/2005, and the 2.3.4.4b lineage strain A/mute_swan/England/117298/2022 viruses, which are currently circulating in wild birds and cattle (Supplementary Figure 3).

Previous studies using live virus assays found that approximately 20% of samples contained low levels of cross-reactive antibodies to 2.3.4.4b lineage H5 viruses [4, 17, 21]. Our data indicate that neutralising antibodies to 2.3.4.4b H5 viruses as well as other H5 viruses, can be detected in a substantially greater proportion of samples than previously demonstrated in live virus neutralisation assays. Our data agree with Daniel et al. [36], with similar findings [4, 40, 39]. We note that our 2.3.4.4b H5 lineage curves (Figure 1A) closely resemble those published by Garretson et al. [17] and Le Sage et al [4] supporting the validity of our data.

The breadth of cross-reactive responses in our cohort extended beyond H5. The H9 A/Hong Kong/308/2014 pseudotyped virus also demonstrated broad population immunity, with over 90% of sera samples showing neutralising activity, displaying age-dependent patterns similar to H5 viruses with peak responses in individuals born before 1957. H9 responses showed strong correlation with H5 viruses (ρ=0.4-0.81), suggesting shared cross-reactive neutralisation mechanisms among Group 1 (H5 and H9) influenza viruses. We also found that 55-65% of samples demonstrated population immunity to potential pandemic H7 viruses A/Shanghai/1/2013 and A/New York/107/2003. Neutralisation levels were generally lower than those observed against seasonal H3 viruses (Supplementary Figure 3). Although this immunity was present at lower levels than H5 cross-reactivity, it demonstrates that the phenomenon of antibody-mediated cross-reactivity to non-circulating influenza viruses is not limited to Group 1 viruses (Figures 1-4). Collectively, these data suggest that there may be low levels of protective antibodies and associated B cells in the human population to potential pandemic H5, H7 and H9 influenza viruses, which could be boosted during an outbreak.

These age-dependent patterns of immunity to Group 1 H5 and H9 subtype viruses are distinct from patterns of immunity to Group 2 H7 subtype viruses, mirroring those identified by Gostic et al [14] and others, in which the incidence and mortality of H5 and H7 influenza infection was highly age-dependent in data derived from Asia. Epidemiological studies by Cowling et al. [22] and Qin et al. [23] have also suggested that older individuals may be more susceptible to H7N9 infection than H5N1 infection. Antibody-mediated population immunity to seasonal viruses also showed clear signs of imprinting, with a 6-year delay between birth year and viral appearance, to which the maximum response is produced. This phenomenon has been reported elsewhere [1, 24, 25] and our data largely align with these studies.

Beyond these age-dependent patterns, we found that IgG1 showed greater binding to H5 antigens compared to circulating seasonal H1 HAs, while IgG2 demonstrated enhanced binding to H7 HA relative to circulating seasonal H3 HAs. Greater IgG1 binding to H5 vs H1 HAs could be explained by greater glycosylation of seasonal H1 HAs, masking epitopes [26].

The higher IgG2 binding observed for A/USSR/90/1977 compared to other H1 HAs may reflect the accumulation of cross-reactive IgG2 responses over decades of repeated seasonal exposure, as this is the most historically distant H1 strain in our panel. Additionally, the USSR HA may harbour fewer glycosylation sites than more recent H1 HAs, exposing protein epitopes more accessible to IgG2 binding [26].

In contrast, the IgG2 dominant response to the H7 HA could be due to Th1 polarised IgG2 class switching over typical IgG1 responses, potentially due to the targeting of subdominant conserved epitopes via repeat H3N2 infection [27, 28]. Whilst IgG3 and IgG4 were present in these samples, the cross-reactivity was mainly mediated by IgG1 and IgG2, which despite having smaller hinge regions were still able to bind to non-circulating and historical seasonal HAs. The low levels of IgG3 could be explained by the lack of recent influenza infection in most blood donors (bloods were collected from healthy donors in March to May 2020, when influenza circulation was comparatively low) and the shorter half-life of IgG3 in comparison to other IgGs.

Via our antibody pulldown assays, we also identified a divergence in head and stem-mediated neutralising immunity to H1 and H5 (Group 1) vs H3 and H7 (Group 2) influenza HAs. H7 was neutralised significantly more by head domain mediated antibodies, whilst H5 viruses were neutralised significantly more by stem domain targeted antibodies. Despite the different proportions of head and stem-targeting neutralising antibodies, neutralising head-targeting antibodies were identified in both instances.

These findings align with previously published studies, many of which corroborate the presence of cross-reactive stem-domain targeting antibodies within Group 1 [8, 17, 29–32] and found that children with primary H3 exposure often had undetectable levels of stem antibodies, while children with primary H1 exposure had low but detectable levels [32]. The observation that H1 infection induces stronger stem antibody responses than H3 infection is supported by Nachbagauer et al. [33], who documented this pattern across all age groups, from young children to the elderly. They hypothesised that sequential infection with antigenically divergent HAs within the same phylogenetic group boosts stem antibody responses. Consequently, the circulation of multiple distinct Group 1 subtypes over the past century may have provided more opportunities to boost stem antibody responses than infection with H3 viruses alone.

Alternatively, the more frequent boosting of H3 versus H1 — due to H3 dominance in three out of five influenza seasons — could provide greater scope for generating broadly reactive head domain antibodies. Notably, Lee et al. found that the head domain of the H3 HA has more mutational tolerance than the stem via deep mutational scanning, which contrasts with the H1 HA variability pattern reported by Doud et al. from the same laboratory, which demonstrated that the head domain of H1 was more intrinsically variable than the stem [34, 35].

We demonstrate that cross-reactive antibodies detected by pMN but not by live virus neutralisation are of significantly lower avidity than those detected by both assays (Figure 5). This avidity-dependent discordance was observed for H3 viruses, where neutralisation is predominantly head-mediated, indicating that the mechanism is not limited to stem-targeting antibodies. Antibodies to the H7 HA were of substantially lower avidity than those targeting the H3 HA, consistent with their generation at greater antigenic distance from the immunising seasonal virus. The lower HA density on pseudotyped compared to live viruses may reduce the number of antibody binding events required for neutralisation, effectively lowering the avidity threshold at which cross-reactive antibodies can be detected. For H5, where neutralisation is predominantly stem-mediated, greater accessibility of stem epitopes on pseudotyped viruses may further contribute to this increased sensitivity. Together, these factors likely account for the variable detection of population immunity to H5 viruses reported across studies using live virus assays [4, 7, 8].Notably, this variability is not unique to comparisons between pseudotype and live virus platforms. Studies using live virus neutralisation assays have reached markedly different conclusions regarding the prevalence of cross-reactive H5N1-neutralising antibodies, with some reporting readily detectable cross-neutralisation in substantial proportions of individuals [4,39,40], whereas others detected little or none [7,8]. These differences likely reflect a combination of cohort characteristics, virus strains tested, and methodological factors that influence the effective avidity threshold required for detection.

Similar discrepancies have been documented in ferret models, where comparable single-infection studies have reported either substantial cross-reactivity to historical H1N1 strains [37] or no detectable cross-reactivity [38]. Together, these observations are consistent with the concept that detection of cross-reactive neutralisation depends not only on the presence of cross-reactive antibodies, but also on whether their functional properties exceed the threshold imposed by a particular assay implementation.

Collectively, our data demonstrate that there is substantial population immunity to potential pandemic viruses, but that this antibody-mediated immunity is divergent in terms of age profile, IgG subclass preference and mediated to different extents by the head and stem of HA. We note that pMN and live virus assay platforms provide complementary information, with the former detecting a broader range of cross-reactive antibody avidities and the latter reflecting the threshold at which neutralisation of replication-competent virus occurs in a cell-based MN assay. Our data support the development of age-specific vaccination approaches and suggest that existing population immunity could potentially be boosted during an outbreak, though protection levels would vary significantly across different age groups.

## Methods Samples

### Blood donor samples

Blood donor samples were obtained from the Scottish National Blood and Transplant Service (SNBTS) collected between 17^th^ of March and 18^th^ of May 2020. Samples were heat-inactivated before serological testing by incubation at 56 °C for 30 min.

Ethical approval was obtained for the SNBTS anonymous archive - IRAS project numbers 18005 and 283057. SNBTS blood donors gave fully informed consent to virological testing, donation was made under the SNBTS Blood Establishment Authorisation and the study was approved by the SNBTS Research and Sample Governance Committee.

### Swan sera

40 sera samples were provided by a previous study focused on understanding the impact of multiple waves of H5 HPAIV infection in a population of mute swans (*Cygnus olor*) on the Fleet Lagoon, Dorset, UK, between 2017 and 2019. All samples were from known individual birds. Most sampled birds are believed based on previous studies to have acquired antibodies to HPAIV. Samples were originally collected under UK Home Office licence P516CDFB6 and with ethical approval from the Animal Welfare and Ethical Review Board at APHA Weybridge for tracking wild bird outbreaks as per [20], but samples used here were residual and therefore no additional ethical permission was required.

### Pseudotyped influenza virus production

Pseudotyped lentiviruses displaying influenza hemagglutinins (HAs) were produced by transfecting HEK 293T/17 cells (ECCAC, Public Health England, UK) with the following plasmid constructs: 1.0 μg of gag/pol construct (p8.91), 1.5 μg of luciferase reporter construct (pCSFLW), 250 ng of TMPRSS4-expressing construct, and 1.0 μg of HA glycoprotein-expressing construct. Transfections were performed in 10 ml of DMEM supplemented with 10% foetal calf serum, 100 U/mL penicillin and 100 µg/mL streptomycin, and 2 mM L-glutamine and incubated for 8 hours at 37°C. To induce virus budding, 1 unit of endogenous neuraminidase (Sigma, USA) was added to 10 ml of fresh media. Culture supernatants were harvested 48 hours post-budding induction, filtered through a 0.45-μm syringe filter, and stored at -80°C. Functional viral entry was confirmed for all pseudotyped viruses by detection of luciferase signal, verifying HA cleavage by TMPRSS4 in each instance.

The influenza strains used for pseudotyped virus production included: A/South Carolina/1/1918 H1N1, A/PR/8/1934 H1N1, A/USSR/90/1977 H1N1, A/Solomon Islands/3/2006 H1N1, A/Brisbane/2/2018 H1N1, A/England/1/1966 H2N2, A/Memphis/1/1968 H3N2, A/Udorn/307/1972 H3N2, A/Netherlands/233/1982 H3N2, A/UK/261/1991 H3N2, A/New York/55/2004 H3N2, A/South Australia/34/2019 H3N2, A/Vietnam/1203/2004 H5N1, A/bar-headed goose/Qinghai/3/2005 H5N1, A/mute swan/England/117298/2022 H5N1, A/Texas/37/2024 H5N1, A/New York/107/2003 H7N2, A/Shanghai/1/2013 H7N9, A/Hong Kong/308/2014 H9N2. The p8.91, pCSFLW, and TMPRSS4-expressing constructs were gifts from Dr. Nigel Temperton. HA-expressing plasmids were produced by cloning GeneArt Strings (Thermo Fisher Scientific, USA) into the pI.18 expression vector (a gift from Dr. Temperton).

### Pseudotyped influenza virus titration

Serial dilutions were made of pseudotyped influenza virus preparations in Corning Costar 96-well plates (Promega, USA). A total of 10^4^ HEK 293 T/17 cells were added to each well and incubated for 3 days at 37 °C. The cells were then lysed with BrightGlo reagent (Promega, USA), and the relative light units of the cell lysate were determined using a Glomax luminometer microplate reader (Promega, USA)

### Pseudotype microneutralisation assay (pMN)

Neutralising antibodies were quantified using a pseudotype microneutralization assay (pMN). Serially diluted sera were added to 96-well Corning Costar plates (Promega, USA) and incubated with 10⁶ relative light unit (RLU) pseudotyped influenza virus for 1 hour at 37°C. Each dilution was performed in duplicate. 6 μL of serum was used per replicate.

Following virus-serum incubation, HEK 293T/17 cells (2.0 × 10⁵ cells/ml) were added to each well and incubated for 3 days at 37°C. Cells were then lysed with BrightGlo reagent (Promega), and relative light units were measured using a Glomax luminometer microplate reader (Promega, USA).

Neutralisation was calculated by comparing RLU values in the presence and absence of antibodies, expressed as percentage neutralisation. The 50% inhibitory concentration (IC₅₀) was defined as the serum concentration that reduced RLU by 50% compared to virus control wells after subtraction of background RLU from cell-only control wells. At least two technical replicates were performed for each biological sample to ensure reproducibility.

### LOWESS

Age-dependent neutralisation data were smoothed using locally weighted scatterplot smoothing (LOWESS), a non-parametric regression method that fits weighted polynomial regressions to localised subsets of data to capture trends without imposing a global parametric model. A smoothing parameter (α) of 0.4 was applied to all curves, meaning that 40% of the data points contributed to each local regression estimate. Confidence intervals (95%) were generated by bootstrapping with 500 iterations, resampling individuals with replacement and recalculating the LOWESS curve for each iteration. All LOWESS analyses were performed using Python 3.12.3.

### Avidity ELISA

Antibody avidity was assessed using a standardised ELISA with a chaotropic wash step to discriminate high- and low-avidity antibodies. NUNC Immuno MaxiSorp 96-well plates (Thermo Fisher Scientific) were coated overnight at room temperature with 50 µL per well of recombinant influenza HA ectodomain at 2 µg/mL in Dulbecco’s PBS. Plates were washed six times with PBS containing 0.05% Tween-20 (PBS/T) and blocked with 200 µL per well of casein blocking buffer (Pierce) for 1 hour at room temperature.

Serum samples were diluted in casein blocking buffer and added in triplicate to coated plates (50 µL per well). Following a 2-hour incubation at room temperature, plates were washed six times with PBS/T. To discriminate antibody avidity, a parallel set of wells received a 10-minute wash with 0.4 M MgCl2 at room temperature, while paired control wells were washed with PBS/T alone. After the chaotropic wash, all wells were washed a further three times with PBS/T.

Bound antibodies were detected by incubation with goat anti-human IgG (γ-chain)-alkaline phosphatase conjugate (Sigma, A3187) diluted 1:1000 in casein blocking buffer for 1 hour at room temperature. Following six washes with PBS/T, plates were developed with 100 µL per well of 4-nitrophenyl phosphate substrate (1 mg/mL in diethanolamine buffer). Absorbance was read at 405 nm using a microplate reader (Promega Glomax). An avidity index was calculated as the ratio of the OD405 after chaotropic wash to the OD405 of the paired control wells, expressed as a percentage.

### Live virus microneutralisation

MDCK-SIAT1 were seeded at 2 × 10⁴ cells per well in 96-well culture-treated plates (Corning) and incubated at 37°C and 5% CO2 overnight. Human sera samples were serially diluted using a 2-fold dilution series in duplicate, starting at a 1:10 dilution, in DMEM supplemented with 0.5% FBS and 2 μg/mL TPCK trypsin (Sigma Aldrich). Virus at 400 TCID50/mL in DMEM supplemented with 0.5% FBS and 2 μg/mL TPCK trypsin was added in equal volume to the diluted sera and incubated for 1 hour at 37°C and 5% CO2. The confluent MDCK-SIAT1 cells were washed with 100 μL PBS, before the diluted sera and virus were added and left to incubate at 37°C for 72 hours. Media was removed, and the cells were stained with Coomassie Blue stain (2% Coomassie Blue powder (Thermo Scientific), 50% ethanol absolute, 7.5% acetic acid, diluted in water) for 30 minutes to assess for cytopathic effect (CPE). Wells with no visible CPE were considered neutralised. On separate plates, the virus was diluted in a 5-fold dilution series and incubated for 1 hour at 37°C and then added to confluent MDCK-SIAT1 cells for 72 hours and checked for CPE to confirm that accurate titres of infectious virus were added. Infectious viral titre was calculated by the improved Spearman-Kärber method to be between 300–500 TCID50/mL. The neutralisation result for a serum sample was determined to be the mean of the highest dilutions showing complete neutralisation of the virus. Positive and negative serum controls were included on each plate. Assay validity was confirmed using ferret antisera raised against the homologous virus where possible.

H3N2 viruses used were: X-31 (A/Puerto Rico/8/1934 (H1N1) reassortant with A/Hong Kong/1/1968 (H3N2) HA and NA), X-79 (A/Puerto Rico/8/1934 (H1N1) reassortant with A/Philippines/2/1982 (H3N2) HA and NA), A/Caen/1/1984 (H3N2) and A/Udorn/307/1972; these assays were performed at BSL-2 at the University of Warwick. H5N1 virus used was A/chicken/England/014330/2022 (clade 2.3.4.4b) and H7N7 virus used was A/chicken/England/1158-11406-1/2008; these assays were performed at BSL-3 at the Animal and Plant Health Agency (APHA-Weybridge). The same microneutralisation protocol was used in both instances. A significance threshold of p = 0.05 was applied to all statistical tests.

### Antibody pulldowns

Neutralising antibodies targeting the HA full-length or head domains of haemagglutinin were purified from donor sera using an ELISA based pull-down method. Viral proteins were added to 96-well MaxiSorp plates (Thermo-Fisher Scientific) at a concentration of 10 μg/mL and incubated overnight at 4°C. Wells were washed with 0.05% PBS-T before wells being blocked with PBS/Casein blocking solution (Thermo-Fisher Scientific) for 1 hour at room temperature. 20uL of donor sera diluted across two wells was incubated for 2 hours at room temperature before removal of non-bound sera was carried out. Elution of bound antibodies was carried out with 3M MgCl2. 5uL of eluted antibodies was used per replicate in a pMN assay as described above. A full protocol is provided in Steventon et al. [42].

### H5 head domain native PAGE

To confirm correct folding of the recombinant H5 HA head domain protein used in antibody pulldown assays, native polyacrylamide gel electrophoresis (PAGE) was performed. The H5 head domain protein was heated at 95°C for 10 minutes to generate a denatured control, alongside an unheated sample. Both heated and unheated samples were separated by native PAGE under non-denaturing conditions. Protein migration was visualised by Coomassie staining. A shift in electrophoretic mobility between heated and unheated samples confirmed the presence of a folded conformation in the unheated protein at the expected molecular weight (Supplementary Figure 11b).

### Structural bioinformatic analysis

Amino acids present on the surface of various H1 HAs were determined by calculating the accessibility of amino acids on the surface of the crystal structures downloaded from RCSB PDB (RCSB.org). Areas of 600, 800 and 1000 Å^2^ were mapped onto the surface of the crystal structures by determining the distances between the α carbon of a given amino acid and all others within a structure. Those residues whose α carbon sequences were within the specified area were recorded and used to produce disrupted peptide sequences for a given binding site. ABS variability was calculated as the mean pairwise hamming distance between the consensus sequences of HA sequences downloaded from GISAID. The sequences were aligned using MUSCLE before being manually curated using AliView.

### IgG subclass analysis via flow cytometry

IgG subclasses specific to viral HAs were characterized using a flow cytometry-based assay. Full influenza HA ectodomains from a panel of eight viruses were used: three H3N2 strains – A/Hong Kong/1/1968 (Stratech, DAG-H10705-CRD), A/Singapore/16/2016 (Stratech, 40580-V08H-SIB), and A/South Australia/34/2019 (Stratech, 40950-V08B-SIB); two H1N1 strains – A/USSR/90/1977 (Stratech, DAG-H10701-CRD) and A/Brisbane/2/2018 (Stratech, 40719-V08H-SIB); one H7N9 strain – A/Shanghai/1/2013 (Stratech, 40104-V08B-SIB); and two H5N1 strains – A/Vietnam/1203/2004 (Stratech, 10003-V06H3-SIB) and A/Texas/37/2024 (Stratech, 41036-V08H-SIB).

Each HA was covalently coupled to 1,000,000 magnetic flow cytometry beads (Spherotech, CMPAK-4068-6K) using carbodiimide chemistry. Beads were activated in 50 mM MES buffer (pH ∼5.0) with 10mg/mL 3-sulfo-N-hydroxysuccinimide (sulpho-NHS; Life Technologies Ltd, 24510) and 10mg/mL 1-Ethyl-3-(3-dimethylaminopropyl)carbodiimide (EDC; Life Technologies Ltd, 22980), then incubated with 5 µg HA antigen for 2 h at room temperature. The beads were then washed and buffer-exchanged using magnetic separation into blocking buffer (PBS with 0.1% BSA [Sigma Aldrich, B6917-100MG] and 0.02% Tween-20 [VWR, MFCD00165986]) and incubated overnight at 4°C to block nonspecific binding. Post-incubation, the beads were distributed evenly into 20 wells of a 96-well U-bottom plate (Corning/Fisher Scientific, 10420862), pelleted, and then resuspended in 50 µL fresh blocking buffer containing 10 µL of individual cross-reactive sera samples. This was allowed to incubate for 1 h at room temperature with gentle agitation.

Following incubation, beads were washed thrice with wash buffer (PBS with 0.1% BSA and 0.05% Tween-20), and then buffer-exchanged into 50 µL blocking buffer containing 2 µg/mL of each fluorescently labelled subclass-specific monoclonal antibody; anti-IgG1-BV421 (BD, 753700), anti-IgG2-PE (BD, 568959), anti-IgG3-R718 (BD, 753748), and anti-IgG4-FITC (Merck, F9890). This incubation was carried out for 1 h at room temperature with gentle agitation.

Beads were again washed thrice and resuspended in 200 µL blocking buffer. The samples were immediately analysed on a flow cytometer (Cytek Aurora for Figure 2 or MACSQuant X for Supplementary Figure 14). Data acquisition aimed for 10,000 events per sample (or a volume of 195 µL to avoid air in the microfluidics), with a minimum of 8,000 events required for inclusion in downstream analysis.

This procedure was repeated for control beads with equal amounts of human IgG1 kappa (Bio-Rad Ltd HCA192), IgG2 kappa (Bio-Rad Ltd HCA193), IgG3 kappa (Bio-Rad Ltd HCA194), and IgG4 kappa (Bio-Rad Ltd HCA195) immobilised on beads. These were used to establish appropriate positive and negative thresholds, as well as calibrate laser gain (Table 1 and Supplementary Figure 8) on the flow cytometer.

Further control beads included three viral antigens (A/Hong Kong/1/1968, A/South Australia/34/2019 and A/Shanghai/1/2013) immobilised on beads, as well as empty control beads, exposed to the secondary antibodies. No cross-reactivity between secondary antibodies and beads or viral antigens was detected. No cross-reactivity between secondary antibodies and non-specific subclasses was detected.

### Deglycosylation of flow cytometry antigens via PNGase treatment

The commercially produced HA ectodomains were deglycosylated to determine the effects of glycans on preferentially bound antibody subclasses, specifically, to discern if the measured IgG2 levels may be attributed to glycan-specific interactions or genuine antibody-protein interactions.

As before, antigens were immobilised on flow cytometry beads. The antigen-containing beads were washed with wash buffer, suspended in 50 µL 1X PBS, and split into two separate aliquots. One aliquot was treated with 1 µL PNGase F (Tebubio, 149101753-1), while the other was treated with 1 µL PBS. Both samples were incubated overnight at 37°C.

The following day, both bead aliquots were pelleted, washed three times with wash buffer, and finally incubated in block buffer for 2 hours. Subsequently, the flow cytometry protocol was followed as previously described. Crucially, the same samples were compared in their response to glycosylated and deglycosylated antigens during the same experimental run, to avoid batch effects arising from, e.g. varying concentrations of secondary antibodies.

### Deglycosylation of pseudotyped influenza via PNGase treatment

The A/Shanghai/1/2013 (H7N9) pseudotyped influenza virus was treated with a deglycosylase enzyme (PNGase F; Tebubio, 149101753-1), to investigate the impact of glycan shields on the neutralisation of pseudotyped influenza viruses.

The pseudotyped influenza viruses synthesised for these purposes were grown following the previously outlined protocol until the media change that accompanies the addition of NA. At this stage, *transfection DMEM* is substituted for serum-free OptiMEM to reduce off-target impurities for the downstream PNGase F treatment. The supernatant was collected 48 hours after the addition of NA. It was centrifuged for 5 minutes at 2,000 × g to pellet any non-adherent HEK293 cells.

The supernatant was concentrated into 1 mL and split into two microcentrifuge vials containing 475 µL aliquots of concentrated supernatant. Both vials were supplemented with 25 µL of 20X PBS, as well as 5 µL of PNGase into one and 5 µL of 1X PBS into the other. Both vials were placed into a 37°C incubator overnight or for at least 10 hours. Following this incubation, both aliquots were briefly incubated with Ni-NTA his-trap beads that were buffer exchanged into PBS. This was aimed at removing the his-tag-containing PNGase. The flow-through was collected, and a 10 µL aliquot was taken from each vial to quality control the deglycosylation via a band shift observed in a western blot.

The remainder of the vials was diluted using Complete DMEM into half its original volume prior to concentration and used in a pMN against 20 samples. Additionally, a broadly neutralising stem antibody (CR8020; Absolute Antibodies, Ab02198) was used to standardise the glycosylated and deglycosylated pseudotyped influenza viruses to one another. As before, these were run in quadruplicate.

### Statistics and Reproducibility

No statistical method was used to predetermine sample size; sample sizes were determined by the volume of archival serum available from the SNBTS blood donor cohort (Supplementary Table 1). No data were excluded. Samples were analysed as anonymised archival sera without experimental intervention; the study was therefore not randomised, and investigators were not blinded to group allocation. All neutralisation assays were performed with at least two technical replicates per biological sample.

Data were assessed before analysis and non-parametric tests were used unless otherwise stated. Mann–Whitney U tests with Holm–Bonferroni correction were used for antibody pulldown (Figure 3, Supplementary Figure 11) and avidity analyses (Figure 5c,d). Site-directed mutagenesis data were analysed by Student’s *t*-test on log10-transformed normalised neutralisation ratios with Holm–Bonferroni correction (Figure 4, Supplementary Figure 12). Wilcoxon signed-rank tests with Bonferroni correction were used for IgG subclass binding analyses (Supplementary Figures 10 and 14). Paired *t*-tests were used for receptor-destroying enzyme treatment (Supplementary Figure 2b), and Spearman’s rank correlation for neutralisation titres (Figure 5a, Supplementary Figures 7 and 13). All tests were two-sided with *P* < 0.05.

Age-dependent neutralisation was visualised using LOWESS (smoothing parameter = 0.4); 95% confidence intervals were estimated by bootstrapping 500 resamples with replacement. Analyses were performed in Python 3.12.3 and GraphPad Prism.

## Data Availability

Source data are provided with this paper. All neutralisation titres, avidity indices, flow cytometry data, antibody pulldown data and site-directed mutagenesis data underlying the figures are provided in the Source Data file and can be found on Figshare [10.6084/m9.figshare.33415582] or GitHub [https://github.com/reannagregory/divergent-immunity-h5-h7-h9]. Influenza virus HA sequences were obtained from the GenBank and GISAID EpiFlu databases (https://www.ncbi.nlm.nih.gov/genbank/, https://www.gisaid.org); accession numbers are listed in Supplementary Table 3. Previously published protein structures used in this study are available from the RCSB Protein Data Bank (https://www.rcsb.org/pages/about-us/index) under accession codes 1RUZ (https://www.rcsb.org/structure/1RUZ), 5E2Y (https://www.rcsb.org/structure/5E2Y), 4WE4 (https://www.rcsb.org/structure/4WE4) and 4LN3 (https://www.rcsb.org/structure/4LN3). Individual-level donor metadata are not publicly available as the sera were obtained under the SNBTS Blood Establishment Authorisation and Research and Sample Governance Committee approval (IRAS 18005 and 283057), which restricts redistribution of donor-linked data. Requests for access to restricted data should be directed to the corresponding author and will require approval from the SNBTS Research and Sample Governance Committee, and access is granted for approved research purposes only; responses can be expected within 28 days. Following approval, restricted data will remain available for a minimum of 10 years from the date of publication.

## Code Availability

Statistical analyses were performed using standard functions in Python 3.12.3 (SciPy v1.14, scikit-learn, statsmodels) and GraphPad Prism [10.2.2]. Code can be found at https://github.com/reannagregory/divergent-immunity-h5-h7-h9 or 10.6084/m9.figshare.33415582.

## Data Availability

All data produced in the present study are available upon reasonable request to the authors post publication and agreement by the SNBTS.

## Acknowledgements

We thank the blood donors who contributed samples through the Scottish National Blood Transfusion Service. We thank Dr Ruth Harvey and Prof Nicola Lewis at the WHO Influenza Centre for providing ferret antisera and influenza strains. We thank Dr Othmar Engelhardt and Dr Basma Bahsoun at the National Institute for Biological Standards and Control (NIBSC) for providing influenza strains.

## Author Contributions

C.P.T. conceived and designed the study. L.S., J.S.B., H.K., R.S., and R.G. performed experiments and analysed data. L.S., J.S.B., and J.H. performed pseudotype microneutralisation assays. R.S. performed antibody pulldown assays and avidity ELISAs. L.S., M.E., and K.C. performed flow cytometry and IgG subclass analysis. M.L. performed live virus microneutralisation assays at Warwick. C.D.G. performed live virus neutralisation assays at APHA. G.C. and N.T. provided pseudotype constructs and reagents, contributed to experimental design and data analysis. L.H., A.B., and J.R. cloned viral constructs. C.B. contributed to experimental design, development and data analysis, and contributed reagents. K.D.C. assisted with pseudotype vector cloning, experimental design, and provided reagents. M.E., T.L., M.C., and P.S. contributed to experimental design and provided reagents. U.O. contributed to data analysis. L.J. and C.M. provided blood donor sera through SNBTS and contributed to experimental design and data analysis. N.C.R. supervised Warwick-based experiments, contributed to experimental design, provided reagents, and helped analyse data. E.M.H. contributed to statistical analysis, data interpretation, and experimental design. S.C.H. provided swan sera and associated data and helped analyse experimental data. A.C.B. and J.J. supervised APHA work, provided reagents, and contributed to experimental design and data interpretation. A.S. and A.M. supervised Oxford-based experiments, contributed to experimental design, and provided reagents. C.P.T. supervised the study and wrote the manuscript with input from all authors.

## Methods Table

**Table 1:** Laser calibration setting on the Cytek Aurora flow cytometer.

| Laser | Gain | IgG subtype |
| --- | --- | --- |
| BV421-A | 142 | IgG1 |
| PE-A | 297 | IgG2 |
| Spark Red 718-A | 233 | IgG3 |
| FITC-A | 342 | IgG4 |

## References

1. Fonville, J.M., et al., Antibody landscapes after influenza virus infection or vaccination. Science, 2014. 346(6212): p. 996–1000.

2. Huang, K.Y., et al., Focused antibody response to influenza linked to antigenic drift. J Clin Invest, 2015. 125(7): p. 2631–45.

3. Allen, J.D. and T.M. Ross, H3N2 influenza viruses in humans: Viral mechanisms, evolution, and evaluation. Hum Vaccin Immunother, 2018. 14(8): p. 1840–1847.

4. Le Sage, V., et al., Influenza A(H5N1) Immune Response among Ferrets with Influenza A(H1N1)pdm09 Immunity. Emerg Infect Dis, 2025. 31(3): p. 477–487.

5. Neumann, G., A.J. Eisfeld, and Y. Kawaoka, Viral factors underlying the pandemic potential of influenza viruses. Microbiol Mol Biol Rev, 2025. 89(2): p. e0006624.

6. Bolton, M.J., et al., IgG3 subclass antibodies recognize antigenically drifted influenza viruses and SARS-CoV-2 variants through efficient bivalent binding. Proc Natl Acad Sci U S A, 2023. 120(35): p. e2216521120..

7. Huang, X., et al., Increase in H5N1 vaccine antibodies confers cross-neutralization of highly pathogenic avian influenza H5N1. Nat Commun, 2025. 16(1): p. 5517.

8. Zhu-Nan Li, F.L., Yu-Jin Jung, Stacie Jefferson, Crystal Holiday, F. Liaini Gross, Wen-Pin Tzeng, Paul Carney, Ashley Kates, Ian A. York, Nasia Safdar, C Todd Davis, James Stevens, Terrence Tumpey, Min Z. Levine, Population Immunity to Hemagglutinin Head, Stalk and Neuraminidase of Highly Pathogenic Avian Influenza 2.3.4.4b A(H5N1) viruses in the United States and the Impact of Seasonal Influenza on A(H5N1) Immunity. Nat Commun 16: 10954 (2025)

9. Wang, J., et al., Broadly Reactive IgG Responses to Heterologous H5 Prime-Boost Influenza Vaccination Are Shaped by Antigenic Relatedness to Priming Strains. mBio, 2021. 12(4): p. e0044921.

10. Gioia, C., et al., Cross-subtype immunity against avian influenza in persons recently vaccinated for influenza. Emerg Infect Dis, 2008. 14(1): p. 121–8.

11. Sanz, I., et al., Heterologous Humoral Response against H5N1, H7N3, and HSN2 Avian Influenza Viruses after Seasonal Vaccination in a European Elderly Population. Vaccines (Basel), 2017. 5(3).

12. Paparoditis, P.C.G., et al., Site-specific serology unveils cross-reactive monoclonal antibodies targeting influenza A hemagglutinin epitopes. Eur J Immunol, 2024. 54(10): p. e2451045.

13. Corti, D., et al., A neutralizing antibody selected from plasma cells that binds to group 1 and group 2 influenza A hemagglutinins. Science, 2011. 333(6044): p. 850–6.

14. Gostic, K.M., et al., Potent protection against H5N1 and H7NS influenza via childhood hemagglutinin imprinting. Science, 2016. 354(6313): p. 722–726.

15. Rimoin, A.W., et al., Ebola Virus Neutralizing Antibodies Detectable in Survivors of theYambuku, Zaire Outbreak 40 Years after Infection. J Infect Dis, 2018. 217(2): p. 223–231.

16. Siber, G.R., et al., Correlation between serum IgG-2 concentrations and the antibody response to bacterial polysaccharide antigens. N Engl J Med, 1980. 303(4): p. 178–82.

17. Garretson, T.A., et al., Immune history shapes human antibody responses to H5N1 influenza viruses. Nat Med 31(5): 1454–1458 (2025)

18. Thompson, C.P., et al., A naturally protective epitope of limited variability as an influenza vaccine target. Nat Commun, 2018. 9(1): p. 3859.

19. Hill, S.C., et al., Impact of host age on viral and bacterial communities in a waterbird population. ISME J, 2023. 17(2): p. 215–226.

20. Hill, S.C., et al., Comparative micro-epidemiology of pathogenic avian influenza virus outbreaks in a wild bird population. Philos Trans R Soc Lond B Biol Sci, 2019. 374(1775): p. 20180259.

21. Writing Committee of the Second World Health Organization Consultation on Clinical Aspects of Human Infection with Avian Influenza, A.V., et al., Update on avian influenza A (H5N1) virus infection in humans. N Engl J Med, 2008. 358(3): p. 261–73.

22. Cowling, B.J., et al., Comparative epidemiology of human infections with avian influenza A H7N9 and H5N1 viruses in China: a population-based study of laboratory-confirmed cases. Lancet, 2013. 382(9887): p. 129–37.

23. Qin, Y., et al., Differences in the Epidemiology of Human Cases of Avian Influenza A(H7N9) and A(H5N1) Viruses Infection. Clin Infect Dis, 2015. 61(4): p. 563–71.

24. Bodewes, R., et al., Prevalence of antibodies against seasonal influenza A and B viruses in children in Netherlands. Clin Vaccine Immunol, 2011. 18(3): p. 469–76.

25. Yang, B., et al., Breadth of influenza A antibody cross-reactivity varies by virus isolation interval and subtype. Nat Microbiol, 2025. 10(7): p. 1711–1722.

26. Altman, M.O., et al., Human Influenza A Virus Hemagglutinin Glycan Evolution Follows a Temporal Pattern to a Glycan Limit. mBio, 2019. 10(2).

27. El-Madhun, A.S., R.J. Cox, and L.R. Haaheim, The effect of age and natural priming on the IgG and IgA subclass responses after parenteral influenza vaccination. J Infect Dis, 1999. 180(4): p. 1356–60.

28. Mai, G., et al., Characterizing the dynamics of BCR repertoire from repeated influenza vaccination. Emerg Microbes Infect, 2023. 12(2): p. 2245931.

29. Ellebedy, A.H., et al., Induction of broadly cross-reactive antibody responses to the influenza HA stem region following H5N1 vaccination in humans. Proc Natl Acad Sci U S A, 2014. 111(36): p. 13133–8.

30. Nachbagauer, R., et al., Induction of broadly reactive anti-hemagglutinin stalk antibodies by an H5N1 vaccine in humans. J Virol, 2014. 88(22): p. 13260–8.

31. Corti, D., et al., Heterosubtypic neutralizing antibodies are produced by individuals immunized with a seasonal influenza vaccine. J Clin Invest, 2010. 120(5): p. 1663–73.

32. Arevalo, C.P., et al., Original antigenic sin priming of influenza virus hemagglutinin stalk antibodies. Proc Natl Acad Sci U S A, 2020. 117(29): p. 17221–17227.

33. Nachbagauer, R., et al., Age Dependence and Isotype Specificity of Influenza Virus Hemagglutinin Stalk-Reactive Antibodies in Humans. mBio, 2016. 7(1): p. e01996–15.

34. Lee, J.M., et al., Deep mutational scanning of hemagglutinin helps predict evolutionary fates of human H3N2 influenza variants. Proc Natl Acad Sci U S A, 2018. 115(35): p. E8276–E8285.

35. Doud, M.B. and J.D. Bloom, Accurate Measurement of the Effects of All Amino-Acid Mutations on Influenza Hemagglutinin. Viruses, 2016. 8(6).

36. Daniel, K., et al., Pre-existing neutralizing antibodies against cattle-transmitted influenza A virus H5N1 are detectable in unexposed individuals. Immunity, 2026.

37. Carter, D.M., et al., Sequential Seasonal H1N1 Influenza Virus Infections Protect Ferrets against Novel 2009 H1N1 Influenza Virus. J Virol, 2013. 87(3): p. 1400–1410.

38. Huang, S.S.H., et al., Immunity toward H1N1 influenza hemagglutinin of historical and contemporary strains suggests protection and vaccine failure. Sci Rep, 2013. 3: 1698.

39. Singh, G., et al., Population immunity to clade 2.3.4.4b H5N1 is dominated by anti-neuraminidase antibodies. mBio, 2026. 17(5): e00445–26.

40. Werner, A.P., et al., Low levels of influenza H5N1 HA and NA antibodies in the human population are boosted by seasonal H1N1 infection but not by H3N2 infection or influenza vaccination. mBio, 2025. 16(12): e02145–25.

41. Bonifaz, L.C., et al., Adjuvanted seasonal influenza vaccination increases cross-reactive antibody responses to highly pathogenic avian influenza H5N1. Nat Commun, 2026. 17(1).

42. Steventon R, et al. Development of an ELISA-based pulldown approach for functional analysis of antigen-specific antibodies. J Virol Methods. 2026 Jul;344:115406.

43. Steventon, R. et al. Antigenic seniority and convergent haemagglutinin evolution shaped the immune landscape preceding influenza B/Yamagata’s extinction. medRxiv (2026). 10.64898/2026.05.17.26353389

