## Supplementary Information for "Divergent antibody-mediated population immunity to H5, H7 and H9 subtype potential pandemic influenza viruses"

### Supplementary Tables


| | |
|---|---|
| **Table 1** | Pseudotyped influenza viruses included in the study, with their corresponding subtype, phylogenetic group, and sample origin. |
| **Table 2** | HPAI live virus neutralisation results. |
| **Table 3** | GenBank and GISAID EpiFlu accession numbers for the haemagglutinin (HA) sequences used to generate the pseudotyped viruses. |


### Supplementary Figures


| | |
|---|---|
| **Figure 1** | Influenza A circulation timeline and phylogenetic tree. |
| **Figure 2** | Pseudovirus microneutralisation assay controls. |
| **Figure 3** | Detectable neutralisation and corresponding $IC_{50}$s against all tested seasonal and potential pandemic viruses. |
| **Figure 4** | Antibody-mediated immune profiles to seasonal influenza strains are age-dependent. |
| **Figure 5** | LOWESS trendlines with bootstrapped 95% confidence intervals for the tested potential pandemic and seasonal influenza strains. |
| **Figure 6** | Antibody-mediated immune profiles for all tested influenza strains separated into 10-year age cohorts. |
| **Figure 7** | Immune profiles of blood donors indicate that neutralisation of one potential pandemic strain is positively associated with neutralisation of other potential pandemic strains. |
| **Figure 8** | Flow cytometry analysis controls. |
| **Figure 9** | Different IgG subclasses preferentially bind to potential pandemic influenza HAs (shared y-axis). |
| **Figure 10** | Effects of deglycosylating HAs or pseudotyped viruses using PNGase results. |
| **Figure 11** | Comparison of head domain and HA1 A/Texas/37/2024 proteins, and H5 HA head domain native PAGE. |
| **Figure 12** | Site-directed mutagenesis of additional H5 and H3 residues, and H5 neutralisation by swan sera. |
| **Figure 13** | H3N2 live virus microneutralisation assay data |

**Figure 14** IgG1 and IgG2 domain targeting of A/Shanghai/1/2013 (H7N9) HA.

**Figure 15** Validation of A/chicken/England/1158-11406-1/2008 as a surrogate for A/Shanghai/1/2013

| Strain Name | Subtype | Origin | Group | Number of blood donors assessed |
|---|---|---|---|---|
| A/South Carolina/1/1918 | H1 | Human | 1 | 337 |
| A/PR/8/1934 | H1 | Human | 1 | 261 |
| A/USSR/90/1977 | H1 | Human | 1 | 260 |
| A/Solomon Islands/3/2006 | H1 | Human | 1 | 253 |
| A/Brisbane/2/2018 | H1 | Human | 1 | 195 |
| A/England/1/1966 | H2 | Human | 1 | 202 |
| A/Memphis/1/1968 | H3 | Human | 2 | 243 |
| A/Udorn/307/1972 | H3 | Human | 2 | 243 |
| A/Netherlands/233/1982 | H3 | Human | 2 | 243 |
| A/UK/261/1991 | H3 | Human | 2 | 243 |
| A/New York/55/2004 | H3 | Human | 2 | 243 |
| A/South Australia/34/2019 | H3 | Human | 2 | 243 |
| A/Viet Nam/1203/2004 | H5 | Human | 1 | 263 |
| A/bar-headed goose/Qinghai/3/2005 | H5 | Avian | 1 | 512 |
| A/mute swan/England/117298/2022 | H5 (2.3.4.4b) | Avian | 1 | 210 |
| A/Texas/37/2024 | H5 (2.3.4.4b) | Human | 1 | N/A |
| A/New York/107/2003 | H7 | Human | 2 | 243 |
| A/Shanghai/1/2013 | H7 | Human | 2 | 243 |
| A/Hong Kong/308/2014 | H9 | Human | 1 | 209 |

**Supplementary Table 1: Pseudotyped influenza viruses included in the study, with their corresponding subtype, phylogenetic group, and sample origin.** Group 1 (H1, H2 and H5 subtypes) viruses assessed individuals from the same 512-member cohort, whilst Group 2 (H3 and H7 subtypes) viruses assessed individuals from the same 243-member cohort.

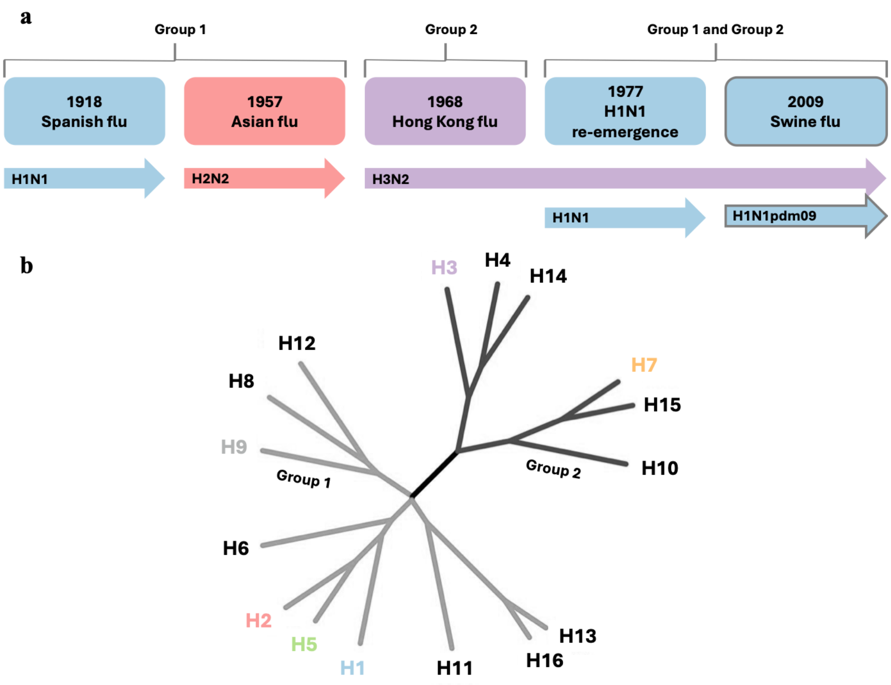


**Supplementary Figure 1: Influenza A circulation timeline and phylogenetic tree.** Figure adapted from *PNAS* 2008. **105**(46): 17736-17741Group 1 HAs consist of H1, H2, H5, H6, H8, H9, H11, H13 and H16. Group 2 HAs consist of H3, H4, H7, H10, H14 and H15.

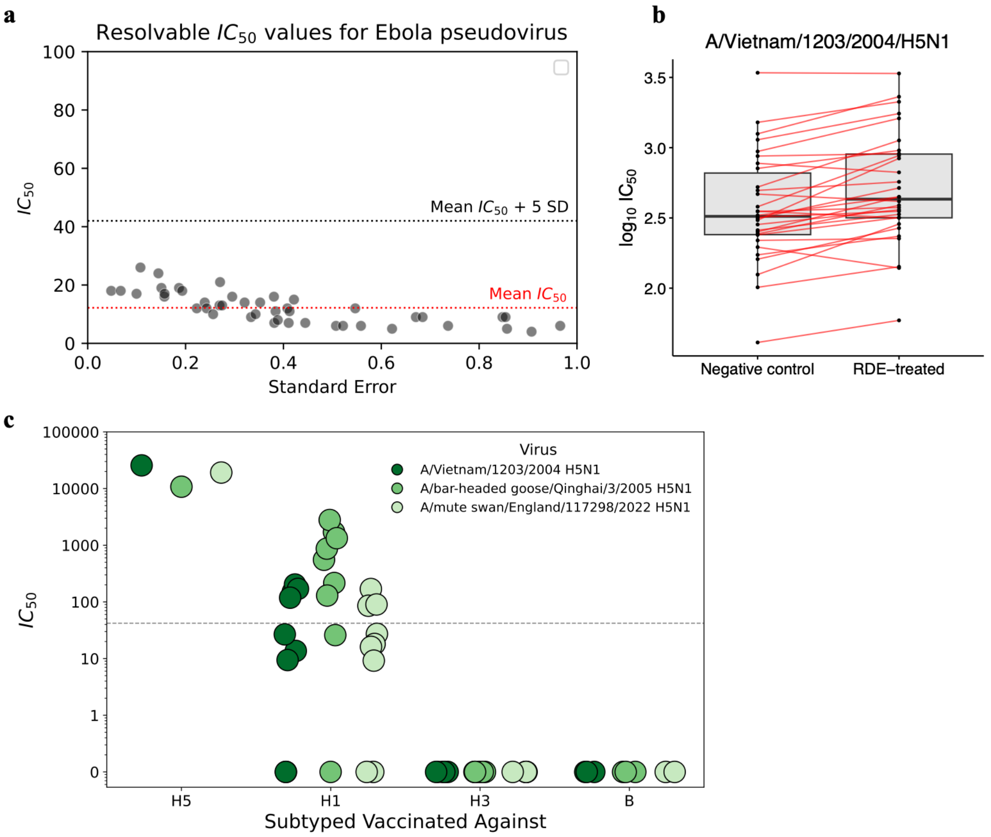


**Supplementary Figure 2: Pseudovirus microneutralisation assay controls. a.** Ebola pseudovirus neutralisation assay. 100 sera samples were run against an Ebola pseudovirus, and a seropositivity threshold was determined as five standard deviations above the mean $IC_{50}$. **b.** Receptor-destroying enzyme assay – 30 sera samples treated with either physiological saline solution or sialic acid-destroying enzyme and used in a neutralisation assay against H5 A/Vietnam/1203/2004 pseudotyped virus. The use of a receptor-destroying enzyme did not remove neutralisation against H5N1; in fact, neutralisation marginally increased. This was statistically significant ($p < 0.001$) according to a paired t-test at a 95% confidence level. **c.** The neutralisation responses of ferrets vaccinated with H1 (n=9), H3 (n=6), H5 (n=1) or Influenza B (n=3) were tested against three H5N1 pseudotyped viruses. Vaccination against H3 or Influenza B did not produce any detectable neutralising antibodies against H5, confirming low assay background. Neutralising antibody responses were greatest in ferrets vaccinated against H5, as expected, with the highest neutralising response against the A/Vietnam/1203/2004 pseudotyped virus that the ferret was specifically vaccinated against. H1 vaccinated ferrets showed detectable neutralising antibodies against H5, that must be due to cross-reactive epitopes between H1 and H5 rather than inherent cross-reactive properties of ferret sera or assay background.

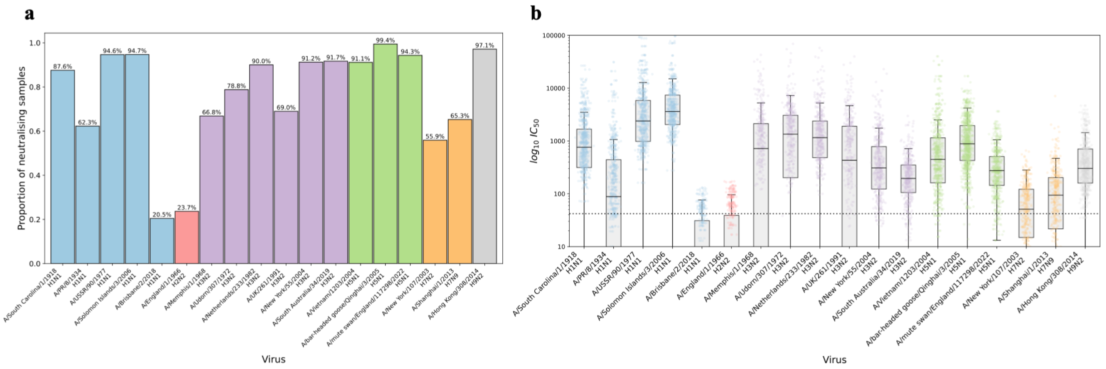


**Supplementary Figure 3: Detectable neutralisation and corresponding $IC_{50}$s against all tested seasonal and potential pandemic viruses. a.** Proportion of neutralising samples for each virus, determined using the seropositivity threshold calculated from the Ebola pseudovirus neutralisation assay (Supplementary Figure S2a). **b.** Corresponding $IC_{50}$s for each virus. Group 1 and Group 2 viruses were run against different sera sample sets from the 2020 blood donor cohort due to sera quantities available. Sample sizes available in Table S1.

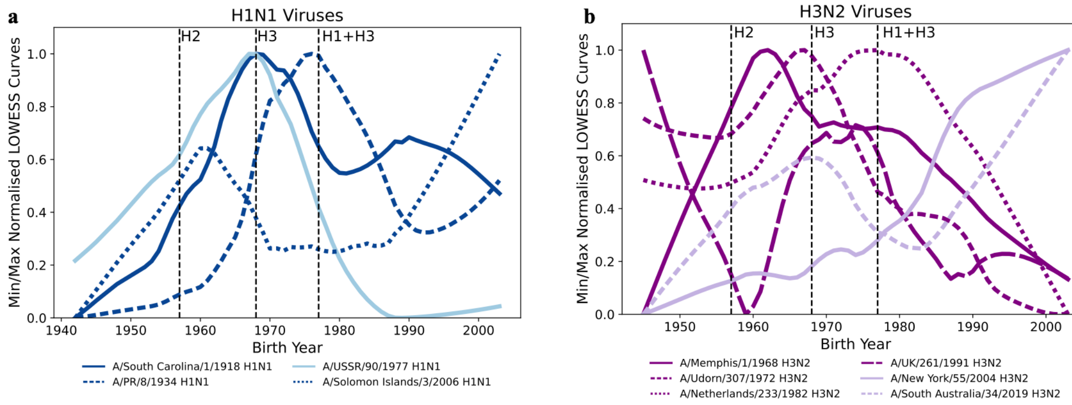


**Supplementary Figure 4: Antibody-mediated immune profiles to seasonal influenza strains are age-dependent. a.** Min/Max normalised LOWESS curves denoting age dependencies for seasonal H1N1 viruses. **b.** Min/Max normalised LOWESS curves denoting age dependencies for seasonal H3N2 viruses. Curves were min/max normalised to allow trends to be directly compared between strains with different $IC_{50}$ ranges. H1 A/Brisbane/2/2018 and H2 A/England/1/1966 pseudotyped virus LOWESS curves were not included as confidence intervals of the line consistently overlapped with zero (see Figure S5). The data was not suitable for min/max normalisation as there were no clear age trends to inflate. Sample sizes available in Table S1.

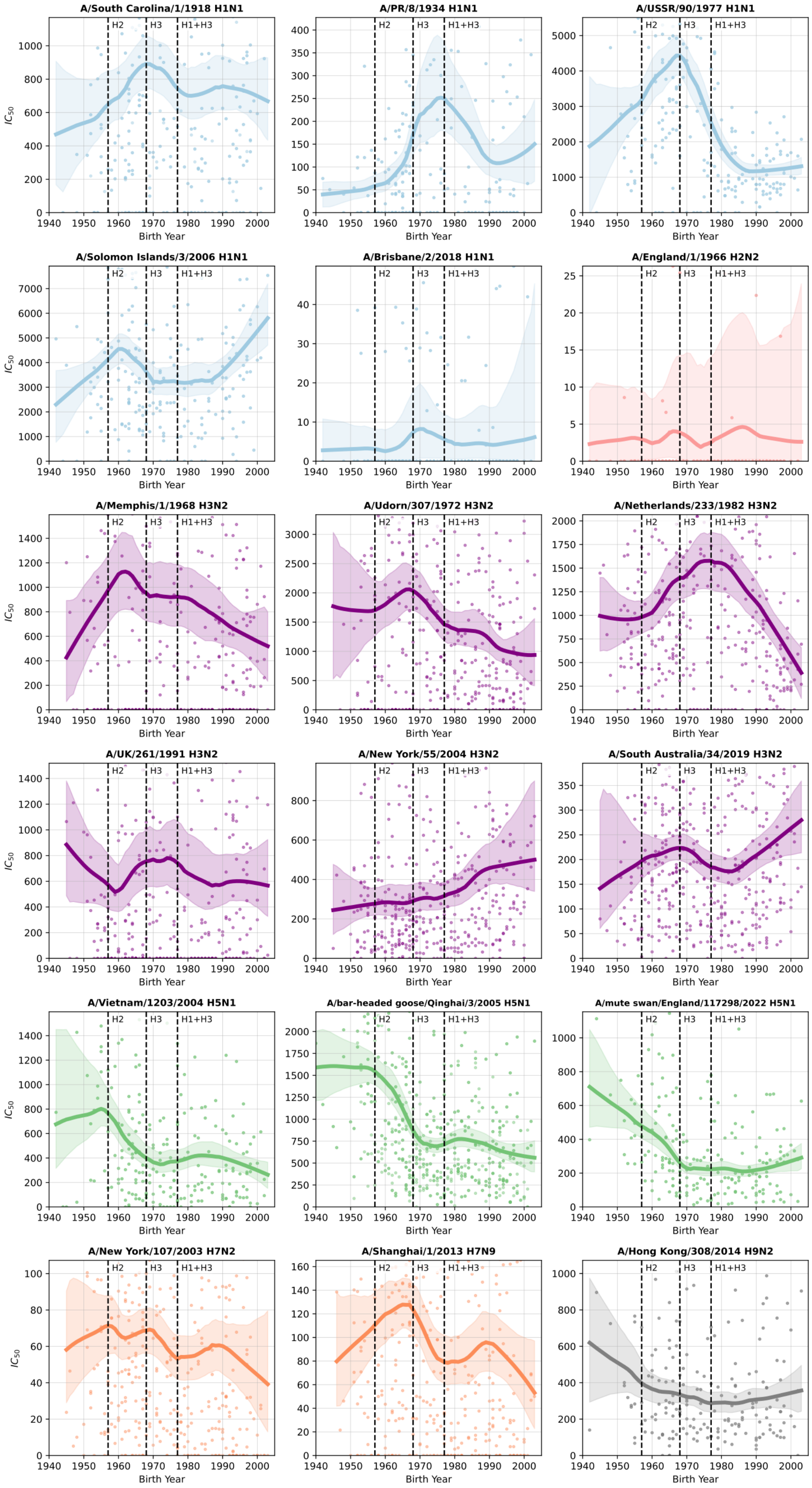


**Supplementary Figure 5: LOWESS trendlines with bootstrapped 95% confidence intervals for the tested potential pandemic and seasonal influenza strains.** A smoothing parameter of 0.4 was used. Sample sizes available in Table S1.

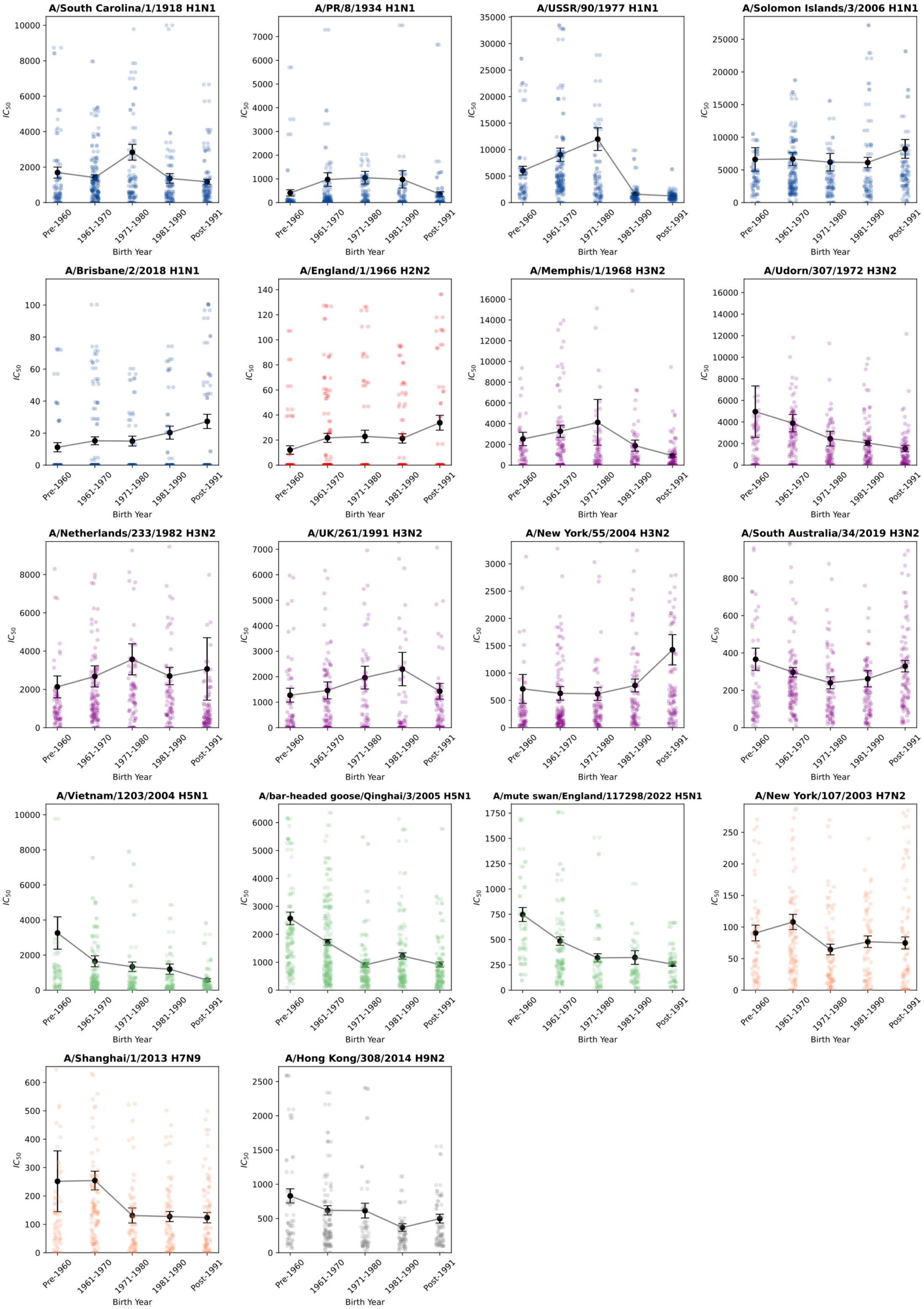


**Supplementary Figure 6: Antibody-mediated immune profiles for all tested influenza strains separated into 10-year age cohorts.** The mean $IC_{50}$ and standard error of the mean (SEM) were calculated for each age cohort. Sample sizes available in Table S1.

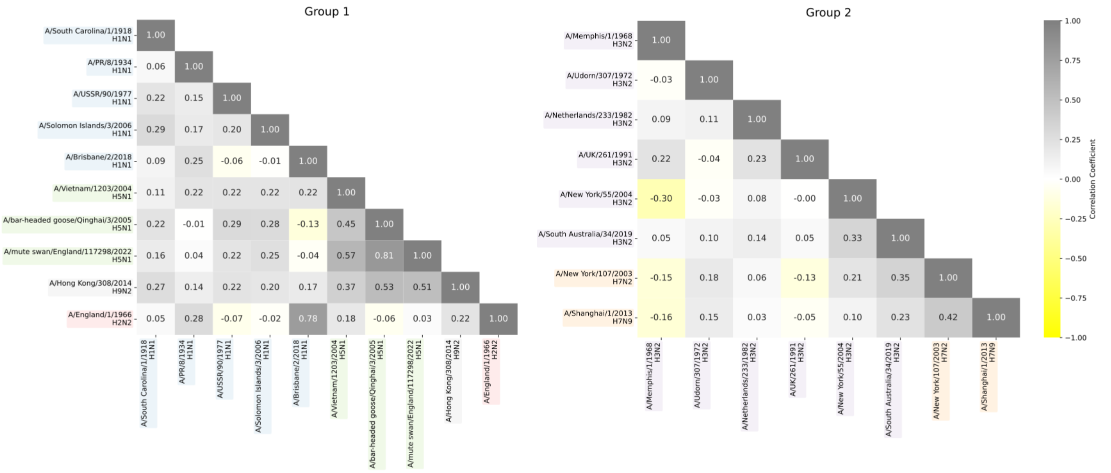


**Supplementary Figure 7: Immune profiles of blood donors indicate that neutralisation of one potential pandemic strain is positively associated with neutralisation of other potential pandemic strains.** Associations were assessed using Spearman's correlation for each virus pair.

**a**

| Antibody | Negative Threshold (a.u.) |
|---|---|
| BV421-*anti*-IgG1 | 1942.74 |
| PE-*anti*-IgG2 | 3110.23 |
| R718-*anti*-IgG3 | 2874.98 |
| FITC-*anti*-IgG4 | 5028.56 |

**b**

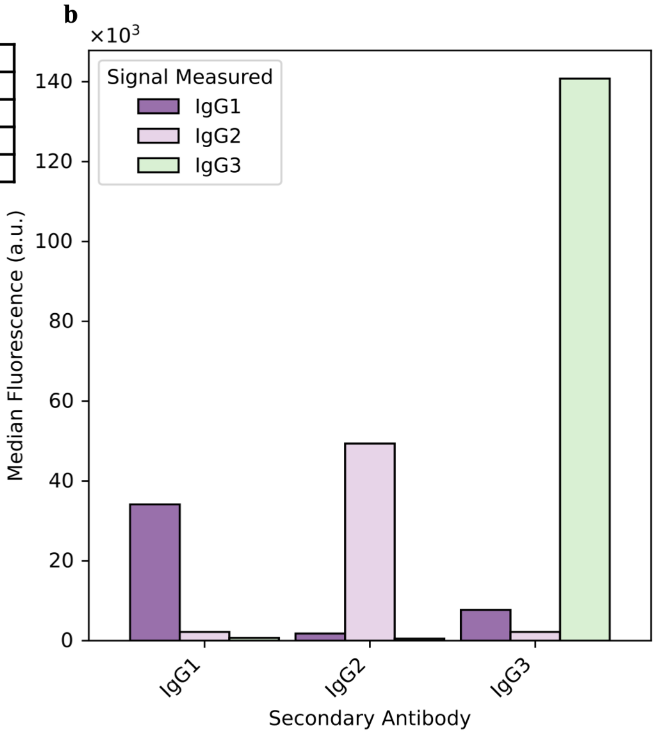


**c**

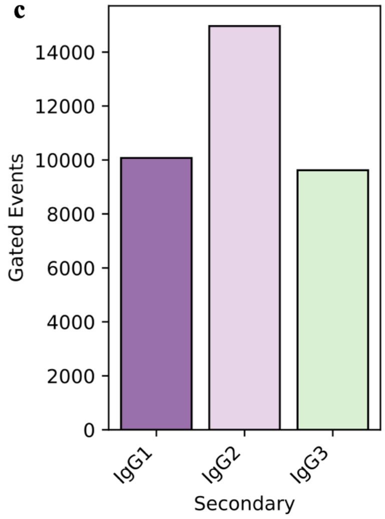


**d**

| Secondary Antibody | Gated Events | IgG1 | IgG2 | IgG3 |
|---|---|---|---|---|
| BV421-*anti*-IgG1 | 10074.00 | 34142.11 | 2192.49 | 699.19 |
| PE-*anti*-IgG2 | 14965.00 | 1783.92 | 49386.00 | 519.06 |
| R718-*anti*-IgG3 | 9620.00 | 7704.51 | 2191.74 | 140795.47 |

**Supplementary Figure 8: Flow cytometry analysis controls. a.** Negative threshold values for each IgG antibody. **b.** The IgG1, IgG2 and IgG3 median fluorescence signals are greatest against the matching secondary antibody. **c.** The number of gated events across IgG1, IgG2 and IgG3 secondary antibodies. All three secondary antibodies achieved similar numbers of gated events. **d.** Supplementary table displaying the raw data used to make panels **b-c**.

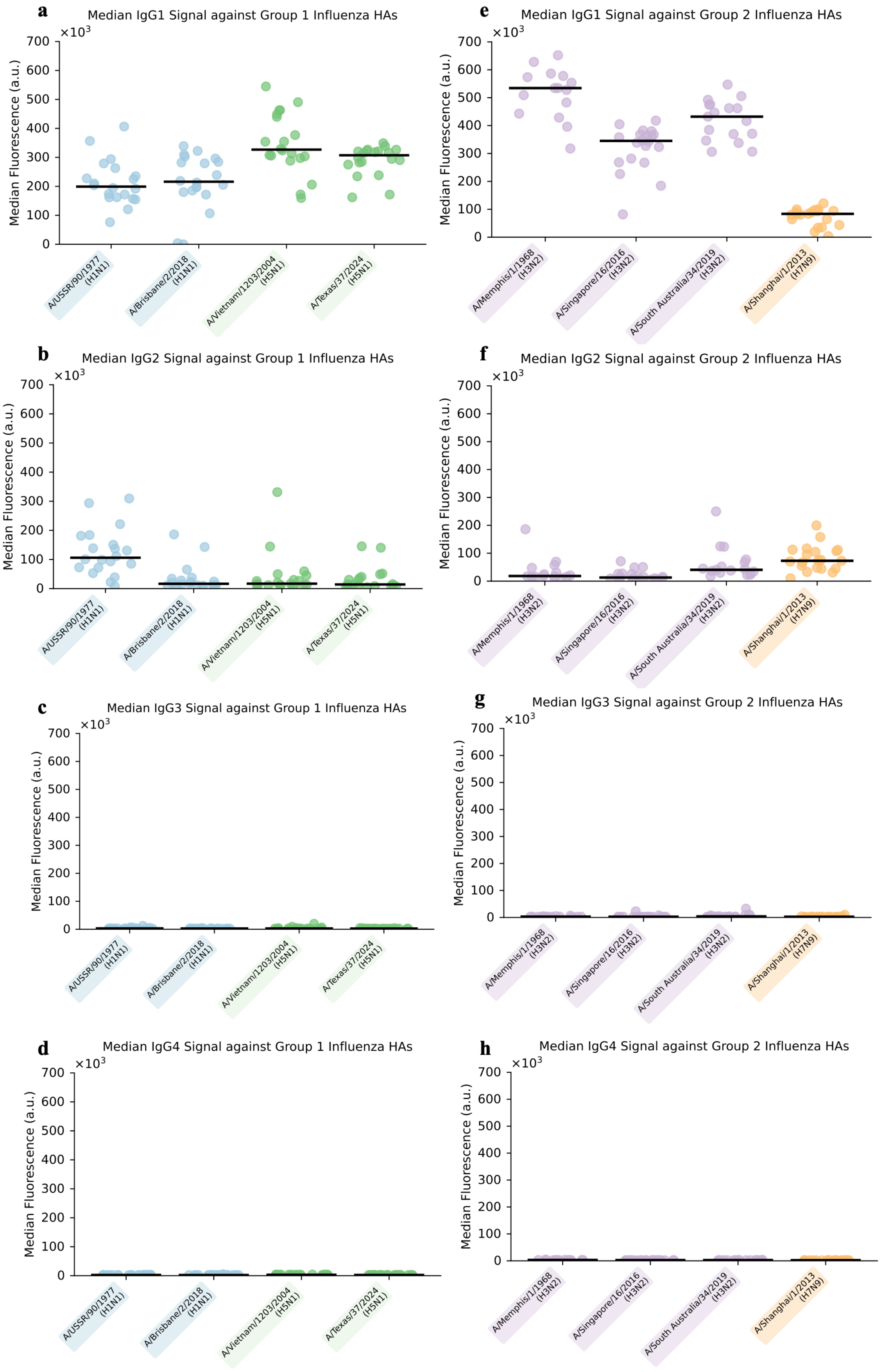
a
Median IgG1 Signal against Group 1 Influenza HAs
b
Median IgG2 Signal against Group 1 Influenza HAs
c
Median IgG3 Signal against Group 1 Influenza HAs
d
Median IgG4 Signal against Group 1 Influenza HAs
e
Median IgG1 Signal against Group 2 Influenza HAs
f
Median IgG2 Signal against Group 2 Influenza HAs
g
Median IgG3 Signal against Group 2 Influenza HAs
h
Median IgG4 Signal against Group 2 Influenza HAs
Median Fluorescence (a.u.)
×10^3
700
600
500
400
300
200
100
0
A/USSR/90/1977 (H1N1)
A/Brisbane/2/2018 (H1N1)
A/Vietnam/1203/2004 (H5N1)
A/Texas/37/2024 (H5N1)
A/Memphis/1/1968 (H3N2)
A/Singapore/16/2016 (H3N2)
A/South Australia/34/2019 (H3N2)
A/Shanghai/1/2013 (H7N9)

**Supplementary Figure 9: Different IgG subclasses preferentially bind to potential pandemic influenza HAs (shared y-axis).** Flow cytometry analysis of IgG subclasses bound to Group 1 and Group 2 full length influenza haemagglutinin (HA), with a shared y-axis across plots to highlight the differences in response magnitude. **a-d**. IgG1-4 composition of Group 1 neutralizing samples run against seasonal H1 – A/USSR/90/1977 and A/Brisbane/2/2018 – and H5 HAs – A/Vietnam/1203/2004 and 2.3.4.4b lineage A/Texas/37/2024. **e-h.** IgG1-4 composition of Group 2 neutralizing samples specifically H3N2 HAs – A/Hong Kong/1/1968, A/Singapore/16/2016, A/South Australia/34/2019 – and H7N9 HAs – A/Shanghai/1/2013.

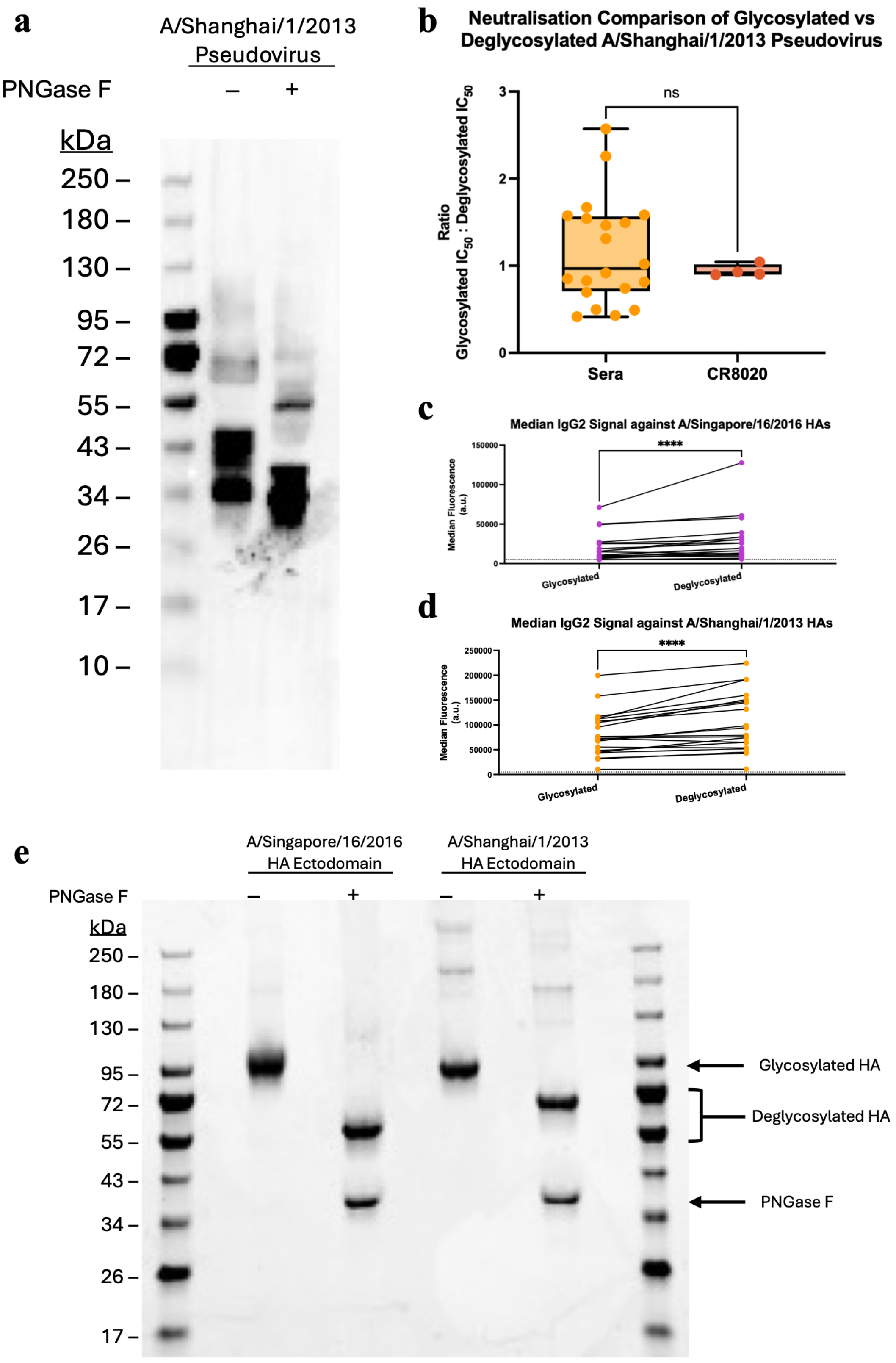
a
A/Shanghai/1/2013
Pseudovirus
PNGase F
–
+
kDa
250 –
180 –
130 –
95 –
72 –
55 –
43 –
34 –
26 –
17 –
10 –
b
Neutralisation Comparison of Glycosylated vs Deglycosylated A/Shanghai/1/2013 Pseudovirus
Ratio
Glycosylated IC50 : Deglycosylated IC50
ns
0
1
2
3
Sera
CR8020
c
Median IgG2 Signal against A/Singapore/16/2016 HAs
Median Fluorescence (a.u.)
****
0
50000
100000
150000
Glycosylated
Deglycosylated
d
Median IgG2 Signal against A/Shanghai/1/2013 HAs
Median Fluorescence (a.u.)
****
0
50000
100000
150000
200000
250000
Glycosylated
Deglycosylated
e
A/Singapore/16/2016
HA Ectodomain
A/Shanghai/1/2013
HA Ectodomain
PNGase F
–
+
–
+
kDa
250 –
180 –
130 –
95 –
72 –
55 –
43 –
34 –
26 –
17 –
Glycosylated HA
Deglycosylated HA
PNGase F

**Supplementary Figure 10. Effects of deglycosylating HAs or pseudotyped viruses using PNGase results. a.** Western Blot band shift showing the effect of deglycosylation on the pseudovirus of A/Shanghai/1/2013. Deglycosylation alters the electrophoretic mobility of HA proteins in polyacrylamide gels by reducing their apparent molecular weight. Accordingly, the PNGase F-treated pseudovirus exhibited a gel shift, migrating further compared to the untreated pseudovirus. A polyclonal primary antibody produced in rabbit (Invitrogen, #PA5-81736) was used. **b.** Effect of glycosylation on the neutralisation of the A/Shanghai/1/2013 pseudovirus. Difference measured by ratio of $IC_{50}$ neutralisation against the glycosylated pseudovirus to the deglycosylated pseudovirus (n=20), when compared to the equivalent ratio achieved by the control antibody, CR8020. Data were not normally distributed; statistical significance was assessed using a non-parametric Kolmogorov–Smirnov test at a 95% confidence interval. **c-d.** Median IgG2 responses for all samples (n=20) across glycosylated and deglycosylated viral HA baits. The data were not normally distributed, so a Wilcoxon matched-pairs signed rank test was performed at a 95% confidence interval. **e.** SDS-PAGE gel shift showing the effect of deglycosylation on A/Singapore/16/2016 and A/Shanghai/1/2013 HA proteins. Deglycosylation alters the migration of HA proteins through a polyacrylamide gel, reflecting changes in molecular weight.

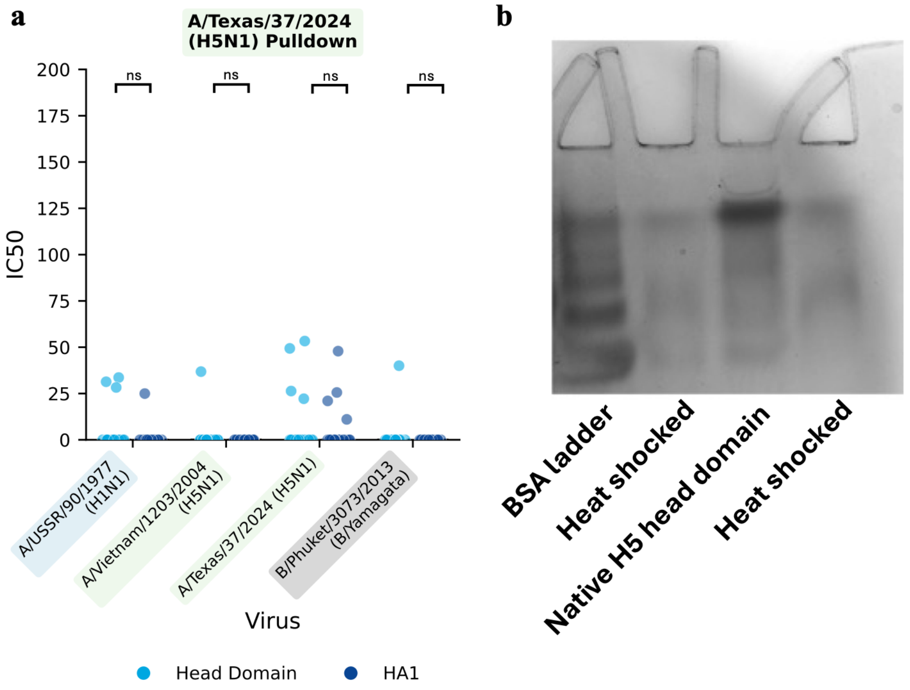


**Supplementary Figure 11: Comparison of head domain and HA1 A/Texas/37/2024 proteins, and H5 HA head domain native PAGE. a.** No statistically significant differences were observed between the head domain or the HA1 proteins (N=20). Statistical analysis was performed using Mann–Whitney U tests with Holm–Bonferroni correction at a 95% confidence level. Asterisks denote statistical significance: $p < 0.05$ (*), $p < 0.01$ (**), $p < 0.001$ (***). **b.** Native PAGE shows the H5 head domain has folded correctly, at the expected size.

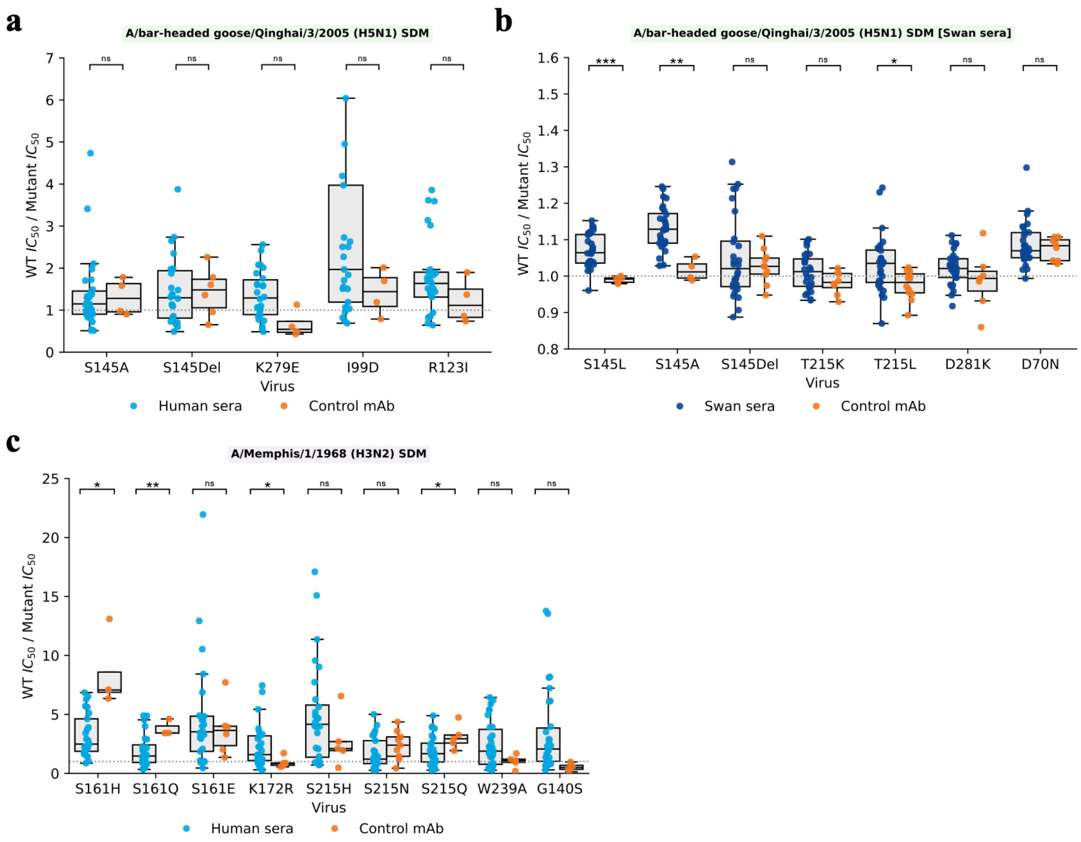


**Supplementary Figure 12: Site-directed mutagenesis of additional H5 and H3 residues, and H5 neutralisation by swan sera. a.** Additional H5 mutations. **b.** H5 mutations tested using sera from swans naturally infected with H5N1. **c.** Additional H3 mutations. Statistical analysis was performed using Student's t-tests with Holm-Bonferroni correction on the $\log_{10}$ of the ratios at a 95% confidence level. Asterisks denote statistical significance: $p < 0.05$ (*), $p < 0.01$ (**), $p < 0.001$ (***). N = 25

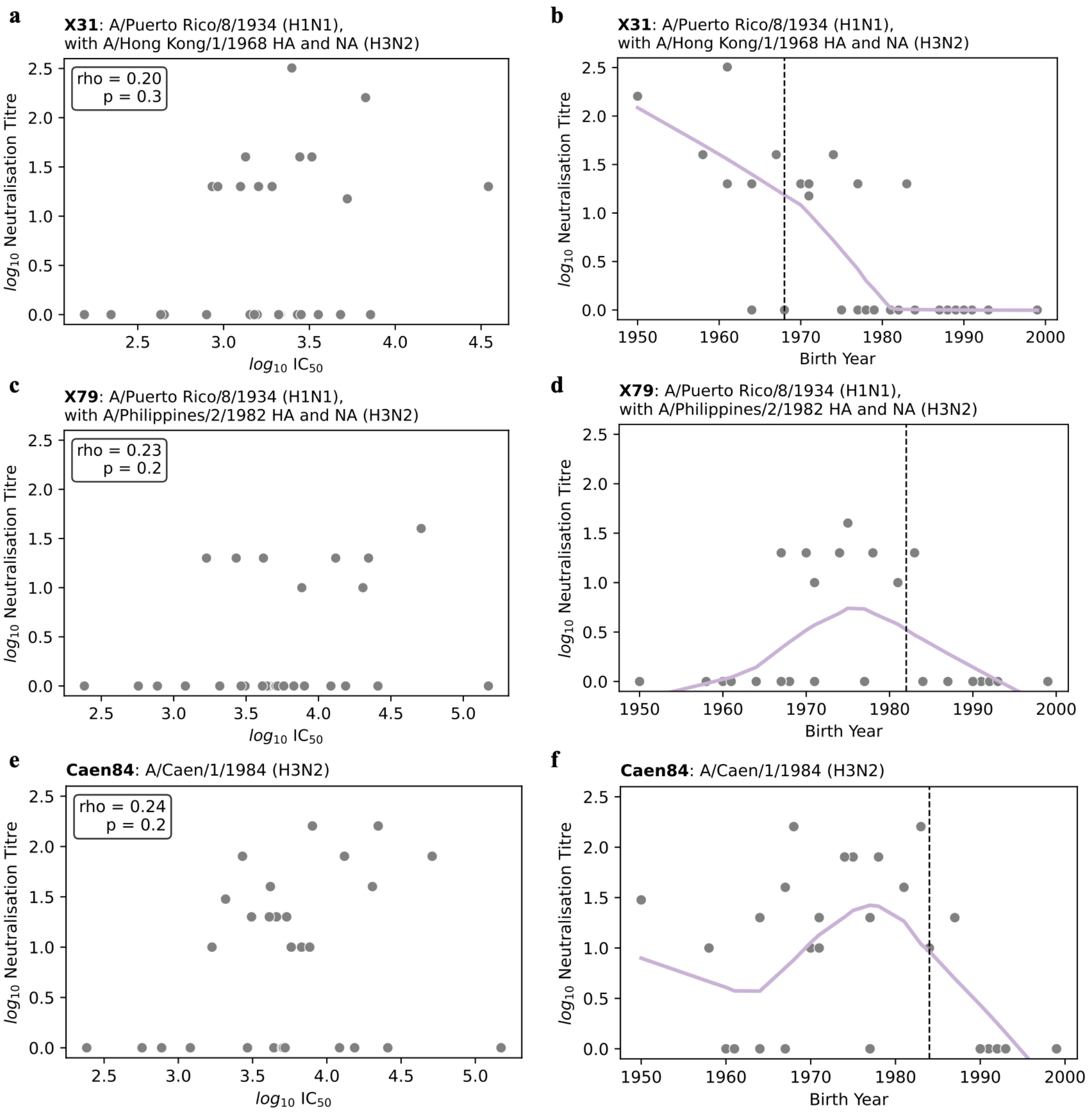


**Supplementary Figure 13: H3N2 live virus microneutralisation assay data. a, c, e.** $Log_{10}$ live virus microneutralisation assay results (neutralisation titre) plotted against pseudovirus neutralisation assay results ($IC_{50}$). Spearman's correlation coefficient (rho) and p-value indicated in top right label. **b, d, f.** $Log_{10}$ live virus microneutralisation assay results (neutralisation titre) plotted against birth year. The vertical dashed line indicates the year that strain began circulating in humans. The greatest live virus titres are primarily found in individuals born prior to the year of viral emergence, indicative of natural infection increasing the likelihood of having detectable antibodies via live virus assay. **a-b.** X31 (A/Puerto Rico/8/1934 (H1N1) reassortant with A/Hong Kong/1/1968 (H3N2) HA and NA), **c-d.** X79 (A/Puerto Rico/8/1934 (H1N1) reassortant with A/Philippines/2/1982 (H3N2) HA and NA), and **e-f.** A/Caen/1/1984 H3N2. 29 samples ran for each virus. LOWESS trendlines are used in panels **b, d, f** to visually indicate trends.

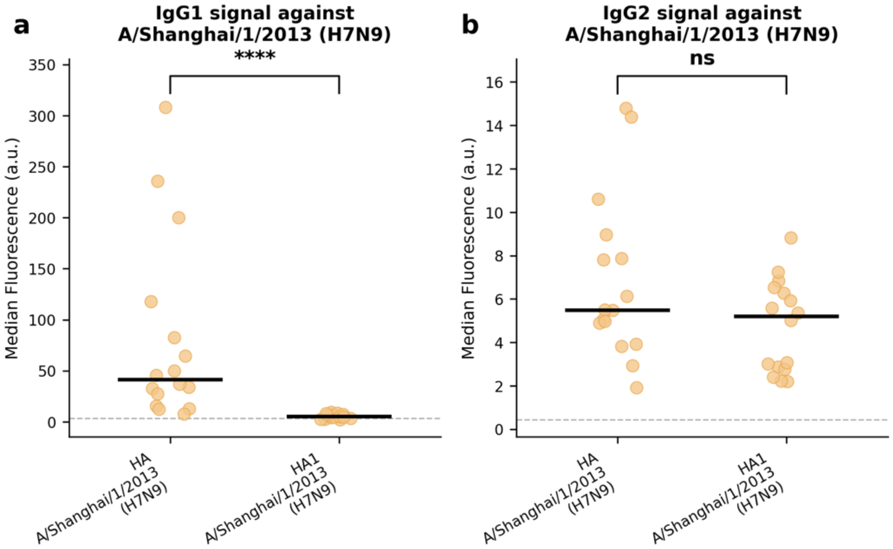


**Supplementary Figure 14. IgG1 and IgG2 domain targeting of A/Shanghai/1/2013 (H7N9) HA.** Flow cytometry analysis of IgG1 and IgG2 binding to full-length HA ectodomain versus HA1 (head domain) of A/Shanghai/1/2013 (H7N9). a. IgG1 binding was significantly reduced when using the HA1 construct compared to the full-length HA (8.0-fold reduction in median GMFI, Wilcoxon $p<0.0001$, adjusted $p<0.001$), confirming that the majority of the IgG1 response targets stem domain epitopes. b. IgG2 binding did not significantly differ between HA1 and full-length constructs (1.1-fold, Wilcoxon $p=0.093$, adjusted $p=0.19$), indicating that the predominant IgG2 response targets head domain epitopes. Horizontal lines indicate medians. Dashed lines indicate the limit of detection based on secondary antibody-only controls. Statistical analysis was performed using Wilcoxon signed-rank tests with Bonferroni correction for multiple comparisons. Compensation was applied for spectral overlap. N = 16. Flow cytometry was performed on a MACSQuant X (Miltenyi Biotec); absolute fluorescence values are not directly comparable between subclasses or to Figure 2 due to differences in instrument calibration.

| Virus | Samples tested | Samples neutralising (%) | Positive control |
|---|---|---|---|
| A/chicken/England/014330/2022 (H5N1 clade 2.3.4.4b) | 30 | 0 (0%) | Infected bird sera positive via HAI assay |
| A/chicken/England/1158-11406-1/2008 (H7N7) | 30 | 0 (0%) | N/A* |

*Virus infectivity confirmed by back-titration. Assay protocol validated using H3N2 viruses with matched positive controls (Figure S13).

**Supplementary Table 2: HPAI live virus neutralisation results.** Live virus microneutralisation was performed at BSL-3 at APHA-Weybridge. No detectable neutralisation was observed in blood donor sera for either virus, despite broad pseudotype neutralisation in the same individuals (93-100% for H5, 55-65% for H7; see Figure 1 and Supplementary Figure S3). Sera positive for A/Shanghai/1/2013 (H7N9) by pseudotype also neutralised pseudotyped A/chicken/England/1158-11406-1/2008 (H7N7), which has 95% sequence identity, confirming cross-reactive H7 antibodies were present in these samples (Figure S15). Virus infectivity was confirmed by back-titration. Assay protocol validated using H3N2 viruses with matched positive controls (Figure S13).

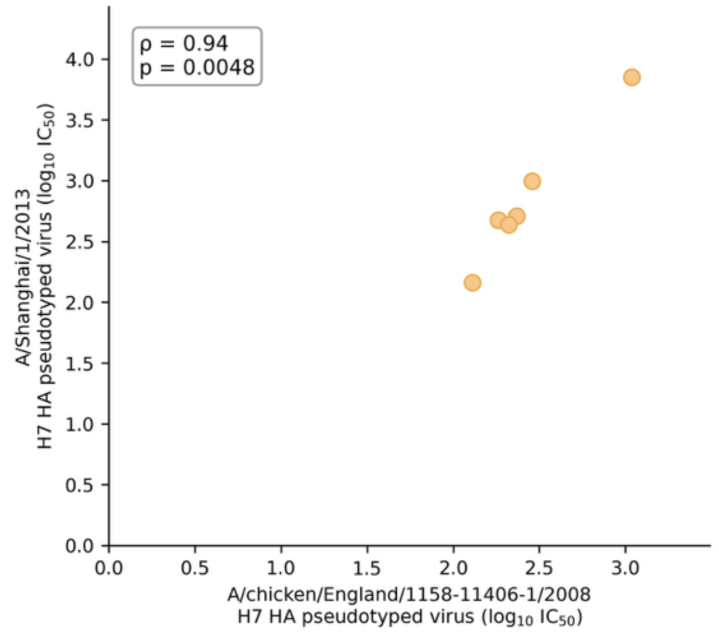


**Supplementary Figure 15. Validation of A/chicken/England/1158-11406-1/2008 (H7N7) as a surrogate for A/Shanghai/1/2013 (H7N9) in live virus neutralisation assays.** Pseudotype microneutralisation IC50 values for A/chicken/England/1158-11406-1/2008 (H7N7) plotted against A/Shanghai/1/2013 (H7N9). Neutralisation titres were strongly correlated (Spearman's $\rho = 0.94$, $p = 0.0048$), supporting the use of A/chicken/England/1158-11406-1/2008 (H7N7) as a representative H7 virus for live virus microneutralisation as A/Shanghai/1/2013 (H7N9) was unavailable. N = 6.

**Supplementary Table 3:** GenBank and GISAID EpiFlu accession numbers for the haemagglutinin (HA) sequences used to generate the pseudotyped influenza viruses in this study.

| Strain name | Subtype | HA accession number |
|---|---|---|
| A/South Carolina/1/1918 | H1N1 | AF117241 (GenBank) |
| A/PR/8/1934 | H1N1 | AF389118 (GenBank) |
| A/USSR/90/1977 | H1N1 | ABF21277.1 (GenBank) |
| A/Solomon Islands/3/2006 | H1N1 | ABU99069 (GenBank) |
| A/Brisbane/2/2018 | H1N1 | WEY08905 (GenBank) |
| A/England/1/1966 | H2N2 | KP412318 (GenBank) |
| A/Memphis/1/1968 | H3N2 | CY006211 (GenBank) |

| | | |
|---|---|---|
| A/Udorn/307/1972 | H3N2 | AAA43099 (GenBank) |
| A/Netherlands/233/1982 | H3N2 | AY661025 (GenBank) |
| A/UK/261/1991 | H3N2 | AAB66755 (GenBank) |
| A/New York/55/2004 | H3N2 | AFM71868 (GenBank) |
| A/South Australia/34/2019 | H3N2 | EPI1387331 (GISAID) |
| A/Viet Nam/1203/2004 | H5N1 | ABW90135 (GenBank) |
| A/bar-headed goose/Qinghai/3/2005 | H5N1 | HM172454 (GenBank) |
| A/mute swan/England/117298/2022 | H5N1 (clade 2.3.4.4b) | EPI2300580 (GISAID) |
| A/Texas/37/2024 | H5N1 (clade 2.3.4.4b) | PP577943.1 (GenBank) |
| A/New York/107/2003 | H7N2 | ACC55270.1 (GenBank) |
| A/Shanghai/1/2013 | H7N9 | KF609511 (GenBank) |
| A/Hong Kong/308/2014 | H9N2 | EPI498034 (GISAID) |
| A/chicken/England/1158-11406-1/2008 | H7N7 | EPI3467395 (GISAID) |
| B/Phuket/3073/2013 | B (Yamagata lineage) | EPI544262 (GISAID) |

### Uncropped gels and blots

*Uncropped scans of all gels and blots shown in the manuscript and supplementary figures. Please indicate on each scan the region reproduced in the corresponding figure panel before submission.*

**Supplementary Figure 10a — A/Shanghai/1/2013 pseudovirus, Western blot (−/+ PNGase F). Uncropped scan.**

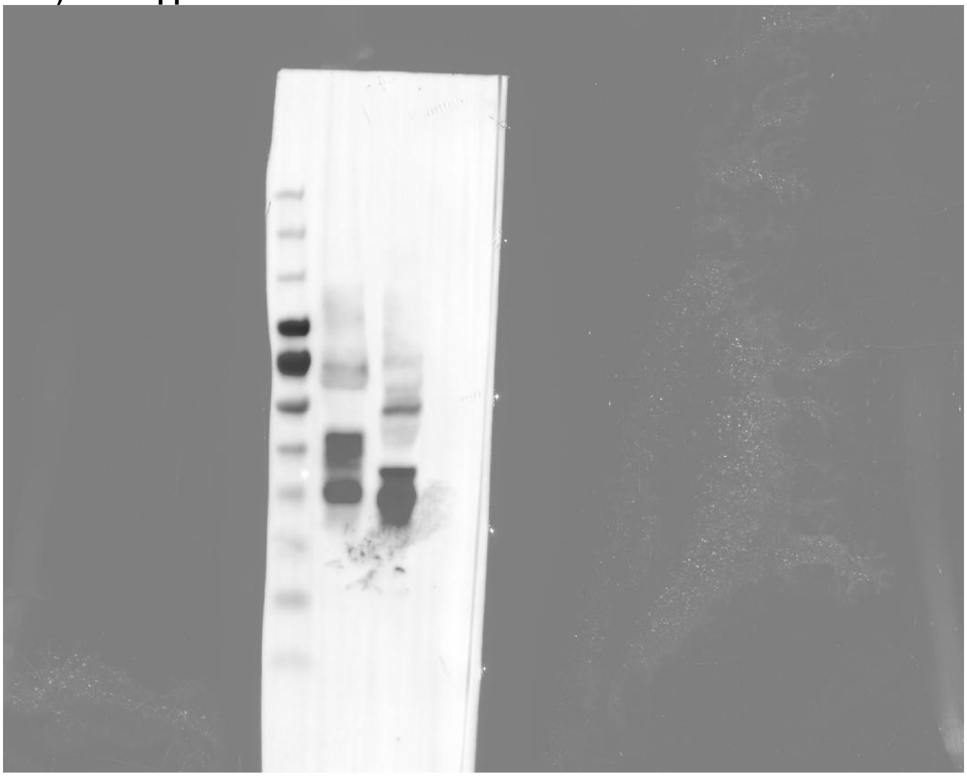

**Supplementary Figure 10e — A/Singapore/16/2016 and A/Shanghai/1/2013 HA ectodomains, SDS-PAGE (−/+ PNGase F). Uncropped scan.**

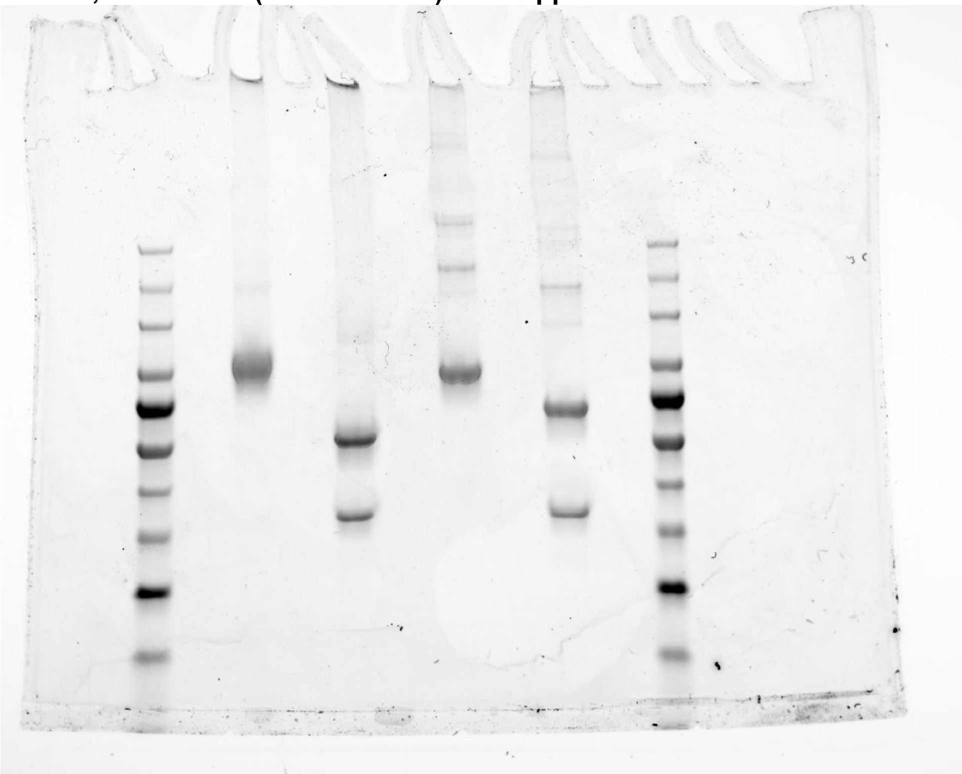

**Supplementary Figure 11b — H5 HA head domain, native PAGE (Coomassie). Uncropped scan.**

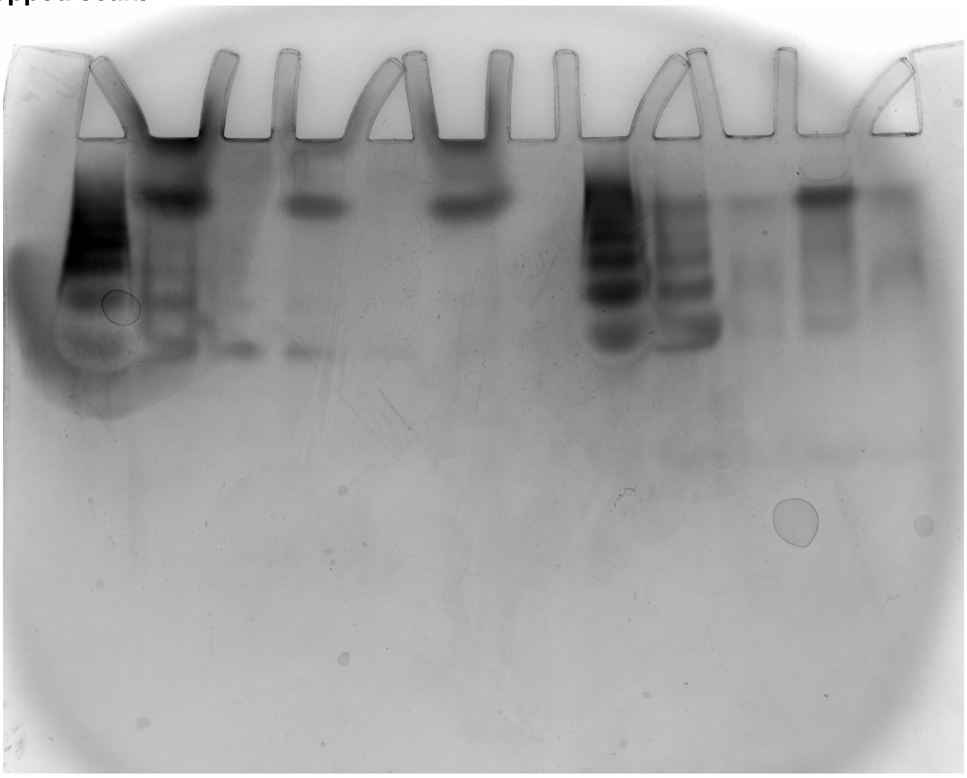
